# Histone H3 lysine 27 acetylation profile undergoes two global shifts in undernourished children and suggests one-carbon metabolite insufficiency

**DOI:** 10.1101/2021.06.11.21258783

**Authors:** Kristyna Kupkova, Savera J. Shetty, Rashidul Haque, William A. Petri, David T. Auble

## Abstract

**Background:** Stunting is a condition in which a child does not reach their full growth potential due to chronic undernutrition. It arises during the first two years of a child’s life and is associated with developmental deficiencies and life-long health problems. Current interventions provide some benefit, but new approaches to prevention and treatment grounded in a molecular understanding of stunting are needed. Epigenetic analyses are critical as they can provide insight into how signals from a poor environment lead to changes in cell function.

**Results:** Here we profiled histone H3 acetylation on lysine 27 (H3K27ac) in peripheral blood mononuclear cells (PBMCs) of 18-week-old and one-year-old children living in an urban slum in Dhaka, Bangladesh. We show that 18-week-old children destined to become stunted have elevated levels of H3K27ac overall, functional analysis of which indicates activation of the immune system and stress response pathways as a primary response to a poor environment with high pathogen load. Conversely, overt stunting at 1-year-of age is associated with globally reduced H3K27ac that is indicative of metabolic rewiring and downregulation of the immune system and DNA repair pathways that are likely secondary responses to chronic exposure to a poor environment with limited nutrients. The results from one-year-old children also point toward deficiency in one-carbon metabolism, which is further supported by integrative analysis with results from histone H3 trimethylation on lysine 4 (H3K4me3).

**Conclusions:** The epigenomes of stunted children undergo two global changes in H3K27ac within their first year of life, which are associated with probable initial hyperactive immune responses followed by reduced metabolic capacity. Limitation of one-carbon metabolites may play a key role in the development of stunting. Trial registration: ClinicalTrials.gov NCT01375647. Registered 17 June 2011, retrospectively registered.

## Background

Stunting is a global health problem in which a child does not reach their linear growth potential due to chronic or recurrent undernutrition. Stunting emerges within the first ∼1,000 days after conception, a crucial time in child development, and if left untreated it can lead to irreversible life-long health problems such as cognitive impairment, a dysfunctional immune system, vaccine failure, and significantly increased risk of mortality below the age of five years (Fig. 1) (1–7). Introduction of a high-fat diet then leads to increased risks of type 2 diabetes, cardiovascular diseases and obesity (8–11). Multiple factors contribute to stunting including nutrient limitation and lack of reliable clean water leading to increased exposure to pathogens. Pathogen exposure often induces episodes of diarrhea, resulting in histopathologic changes to the intestine and delayed or failed microbiome maturation (9,12,13). Additional contributors to stunting include socioeconomic status, maternal health, and other environmental variables such as toxins (1–3). In 2020, it was estimated that 21.3% (144 million) children under the age of 5 worldwide were stunted (5), with higher rates in low- and middle-income countries. The issue is so prominent that the World Health Organization (WHO) has set a goal to decrease the rate of stunting by 40% by 2025 (14). To accomplish this, new nutritional interventions are needed, particularly those provided to children living in urban slum areas of low- and middle-income countries (7,15).

**Fig. 1.**
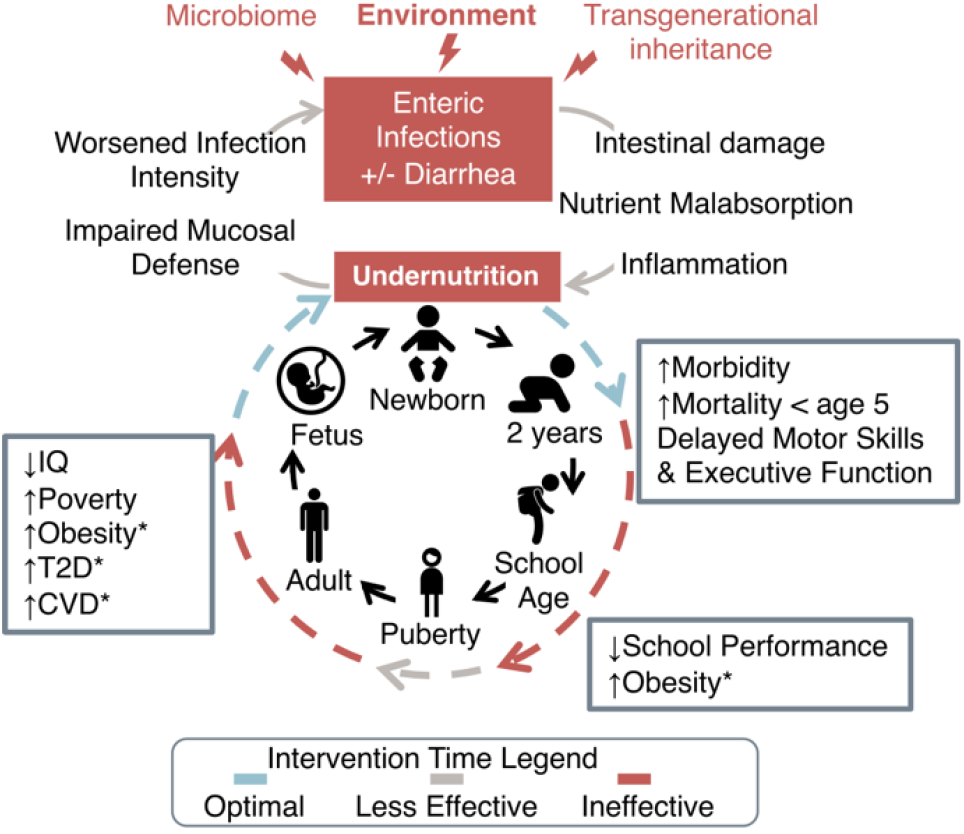
Vicious cycle of chronic undernutrition. Multiple factors contribute to chronic undernutrition in children. Enteric infections (with or without diarrhea) are key determinants of a child’s nutritional status, as they often lead to gut dysfunction with accompanying health issues listed in the figure. An optimal time for intervention is from conception until 2 years of age, a crucial time in child’s development (2). A stunted child faces a high risk of life-long health consequences, which may be passed on to the next generation through poorly understood mechanisms. Health problems associated with the introduction of high calorie diet are marked with an asterisk. T2D -type 2 diabetes, CVD – cardiovascular diseases.

We reasoned that the identification of epigenetic differences associated with stunted children would shed light on how the gene regulatory circuitry is modified in response to a poor environment. The epigenetic state of a cell is tightly linked to the availability of key metabolites (16), which makes it a fundamentally important area of focus in relationship to stunting. In our previous work we showed that H3 trimethylation on lysine 4 (H3K4me3) profiles of stunted children undergo large-scale changes within the first year of life, which are similar to changes observed in cells grown in low methionine (17). Here we expand on this finding by analyzing histone H3 acetylation on lysine 27 (H3K27ac). H3K27ac provides an indication of the relative activation state of transcriptional regulatory regions, particularly enhancers (18). The discrete temporal changes in H3K27ac that we observe as stunting unfolds point to hyperactive immune responses in early infancy followed by metabolic downregulation by the time stunting is evident at one year of age. These patterns provide a framework for future development of biomarkers for at-risk infants and potentially new nutritional interventions to reset metabolic function, including one carbon metabolism.

## Results

### Profiling the H3K27ac landscape in stunted and healthy infants

We obtained samples of peripheral blood mononuclear cells (PBMC) from 18-week-old and 1-year-old Bangladeshi infants enrolled in the PROVIDE (“performance of rotavirus and oral polio vaccines in developing countries”) study (19) and performed chromatin immunoprecipitation followed by sequencing (ChIP-seq) to profile histone H3 acetylation on lysine 27 (H3K27ac) (Fig. 2a). We chose H3K27ac, a histone marks associated with active regulatory elements, as it has been shown to distinguish active from poised enhancers, and due to its roles in development (18,20). Stunting emerges within the first two years of life, and for this reason the first two years are considered an optimal time for therapeutic interventions. A child is considered stunted when his/her height-for-age z-score (HAZ) is below -2 by one year of age. The available anthropometric data included each child’s HAZ scores at birth, 18 weeks, and 1 year (Fig. 2a, Additional file 1: Table S1), which allowed us to track epigenetic changes scaling with growth trajectory (the change in HAZ score over time, ΔHAZ), rather than simply using the phenotypic classification at a given point in time. In our prior work, we discovered that the analysis of epigenetic changes using growth trajectory afforded a much richer picture of chromatin changes than using phenotypic classification or HAZ scores alone (17). The results obtained using ΔHAZ are more informative about factors associated with the development of stunting because the HAZ score often changes dynamically during early infancy, and children born with low HAZ scores are not certain to becoming stunted, and vice versa (Fig. 2a). Thus, the context provided by using ΔHAZ allows for more accurate and meaningful analysis.

**Fig. 2.**
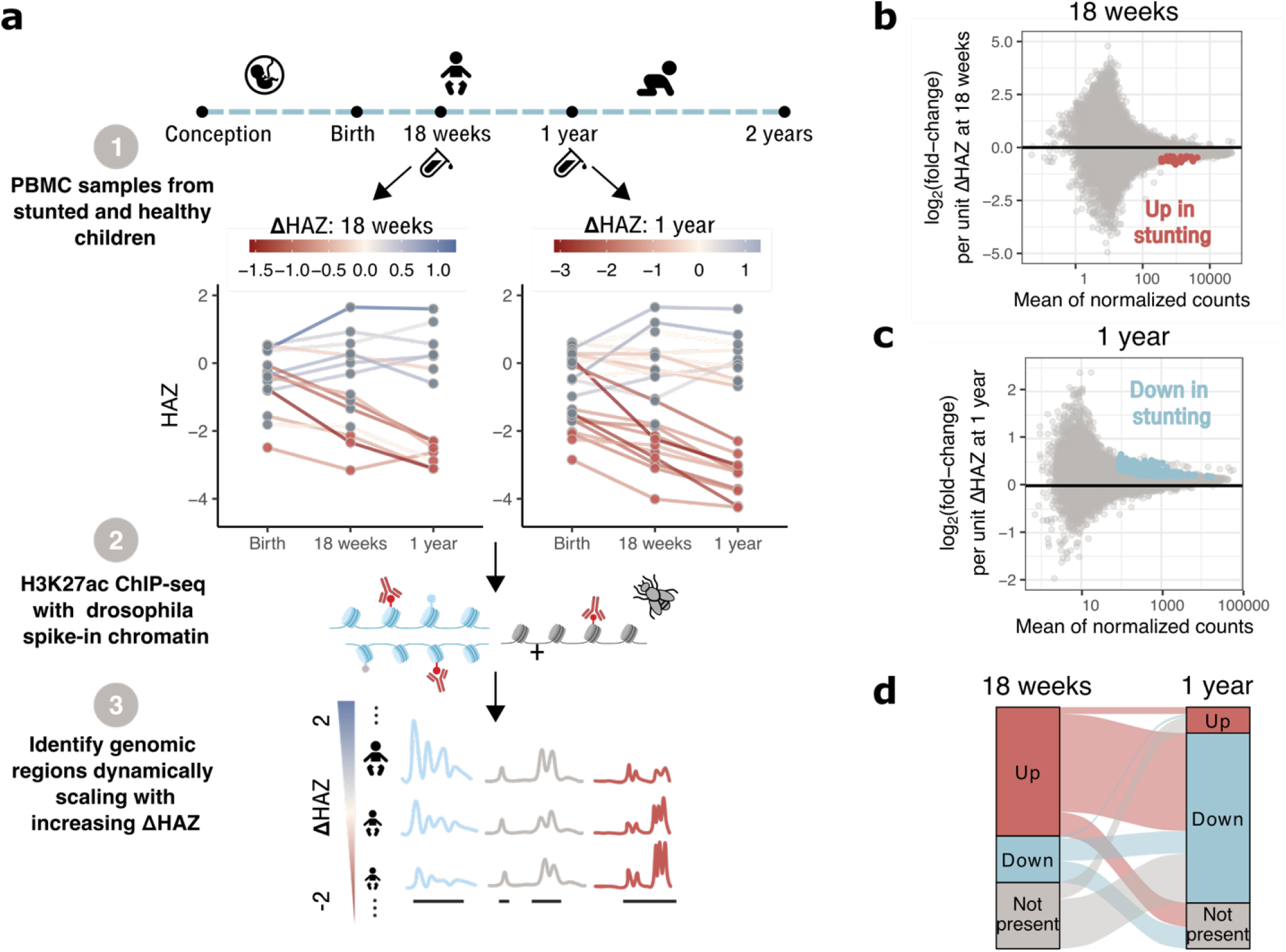
The H3K27ac landscape undergoes global changes in stunted children within their first year of life. (a) Overview of the experimental design. Each line in the plot with HAZ scores represents the change of HAZ score from birth to 1 year of age for a child whose PBMC sample was used for the study. The left panel shows HAZ scores for 18-week-old children, the right panel for 1-year-old children. Red dots indicate HAZ < -2 at a given age. Lines are colored based on ΔHAZ between birth and 18 weeks or between birth and 1 year. Bottom panel illustrates three different scenarios for differential analysis in which blue regions are downregulated in stunting, grey regions unchanged or with no significant trend; and red regions are upregulated in stunted children. (b) MA-plot shows changes in H3K27ac regions in 18-week-old children as the ΔHAZ (18 wk) score increases. Each dot is an H3K27ac region, the x-axis represents the mean read coverage over the region, and the y-axis indicates the log_2_(fold-change) of read coverage per unit increase of ΔHAZ (18 wk) score. Colored dots indicate significantly affected regions with false discovery rate (FDR) corrected p-value < 0.05. (c) Same as (b) for 1-year-old children. (d) Alluvial plot showing changes to log_2_(fold-change) of corresponding regions between 18 weeks and 1 year. “Up” / “down” – H3K27ac levels are increased / decreased, accordingly, with higher risk of stunting at a given age, “not-present” – region was not identified at the given age.

The samples came from both male and female infants and with a broad range of HAZ and ΔHAZ scores (Additional file 2: Fig. S1a-c, j-l, Additional file 1: Table S1). Initial analyses did not suggest any differences in the number of H3K27ac enriched regions (peaks), putative enhancers, or superenhancers in association with any of the anthropometric z-scores (Additional file 2: Fig. S1d-i, m-r, Additional file 3: Table S2). We therefore moved on to analysis of H3K27ac levels in enhancer elements and their relationships with ΔHAZ scores.

### H3K27ac profile undergoes two global shifts in stunted children within the first year of life

We added *Drosophila melanogaster* chromatin as a spike-in control to determine if H3K27ac levels were globally altered among the samples (Fig. 2a, Additional file 2: Fig. S2, Additional file 3: Table S2). Principal component analysis (PCA) carried out on normalized H3K27ac counts shows a gradient change in the first two principal components (PCs) associated with ΔHAZ scores, where healthy children tend to locate in the upper right corner of the PCA plot and gradually progress to the lower left quadrant as the ΔHAZ score decreases (Additional file 2: Fig. S3a,b). The gradient is more apparent along PC1 and is especially pronounced in 18-week-old children. These results suggest that there are changes to H3K27ac levels associated with stunting in both 18-week-old and 1-year-old children.

Results of differential analysis versus ΔHAZ scores while controlling for sex indicated 38 significantly upregulated regions in 18-week-old stunted children (Fig. 2b, Additional file 4: Table S3), and 341 significantly downregulated regions in 1-year-old stunted children (Fig. 2c, Additional file 5: Table S4), the biological roles of which are discussed below. In addition to the significantly affected regions, we observed a significant global shift in H3K27ac indicating increased acetylation in 18-week-old stunted children (Fig. 2b, Additional file 2: Fig. S3c) and globally decreased acetylation at 1 year of age in stunted children (Fig. 2c, Additional file 2: Fig. S3d) compared to control children with higher ΔHAZ scores. This indicates that for children destined to become stunted, the H3K27ac landscape undergoes two major global shifts within the first year of life. Both shifts are statistically significant (p = 4.1 · 10^−196^, and p = 0 for 18-week-old children and 1-year-old children, respectively; Additional file 2: Fig. S3c,d). We also observed global shifts in the same directions in putative superenhancer regions, none of which, however, showed statistically significant association with ΔHAZ score (Additional file 2: Fig. S3e,f).

To test whether the observed patterns in the H3K27ac data might be driven by a specific subpopulation of PBMCs, we mapped the normalized H3K27ac signal onto cell type-specific enhancers defined by Andersson et al. (2014) (21), and calculated the average H3K27ac profile for each cell type (Additional file 2: Fig. S4a). If H3K27ac changes were attributable to a specific blood cell subtype, we would expect this particular subtype to show a pattern in which the average profile height would be positively or negatively correlated with the ΔHAZ scores. As shown in Additional file 2: Fig. S4b,c, the ordering of average profiles was similar across all the different cell types, suggesting that the shifts that we observe in stunted children are not indicative of changes in a specific population of blood cells compared to the others.

### Changes in H3K27ac between 18-weeks and 1-year of age

We matched enhancer regions from 18-week-old and 1-year-old children and compared the differential H3K27ac results directly to identify changes in enhancer acetylation that occur during this period of developmental time (Additional file 2: Fig. S5a). As a complementary strategy, we classified the differential acetylation results into three categories: 1) “up” – regions that were upregulated in stunting, 2) “down” – regions that were downregulated in stunting, and 3) “not present” – regions that were not detected at a given age (Fig. 2d). As expected based on the results that we have already presented, the largest proportion of regions were upregulated in 18-week-old stunted children and downregulated in 1-year-old stunted children (referred to as “up-down” Fig. 2d, Additional file 2: Fig. S5a,b). The second largest group consists of regions that were not detected in 18-week-old children and were downregulated in 1-year-old stunted children (referred to as “not present-down”; Fig. 2d, Additional file 2: Fig. S5b). Interestingly, even though the “up-down” regions were the dominant group of changes between 18 weeks and 1 year, we did not observe any correlation between the changes associated with stunting at 18 weeks and 1 year of age (Additional file 2: Fig. S5a).

Next, we assessed the functional significance of regions in the two largest identified groups: 1) “up-down,” and 2) “not present-down,” by assigning the regions to their target genes followed by functional enrichment analysis of the associated genes. As shown in Additional file 2: Fig. S5c, clear patterns of pathway enrichment were observed. Genes with enhancers in the “up-down” category were associated with pathways involved in signaling processes, mRNA splicing, transcription-coupled nucleotide excision repair (TC-NER), transcription, translation, and the general stress response. These results suggest that infants destined to become stunted are not able to sustain responses to DNA damage and other stresses resulting from long term exposure to a detrimental environment. Since TC-NER is activated by DNA damage in order to prevent cytotoxic transcriptional stress (22), TC-NER perturbations are related to deficits in transcription and associated pathways. The genes associated with “not present – down” regions show enrichment in immunological pathways, which suggests that the immune system of children with a high risk of stunting does not reach its full developmental potential.

### Gene targets of upregulated H3K27ac regions in 18-week-old stunted children

We next analyzed the 38 H3K27ac regions that showed significant association between H3K27ac levels and ΔHAZ (18 wk) scores (Fig. 2b). All of the regions show increased coverage with decreasing ΔHAZ (18 wk) score, indicating that H3K27ac levels progressively increased with the degree of stunting. An example region is shown in Fig. 3a. Functional annotation of target genes (Fig. 3b) identified pathways involved in TC-NER, transcription, and viral infection. Even though HIV infection pathways were seen to be enriched, further enrichment analysis of the genes found in these particular pathways showed that these genes are actually involved in general viral response, as shown in Fig. 3c. These results suggest that children destined to become stunted face higher pathogen loads, possibly resulting from intestinal damage due to prior enteric infections (23), which additionally lead to increased DNA damage (24) among other associated problems. Eighteen-week-old infants may then activate the aforementioned pathways in response to these pathogen challenges.

**Fig. 3.**
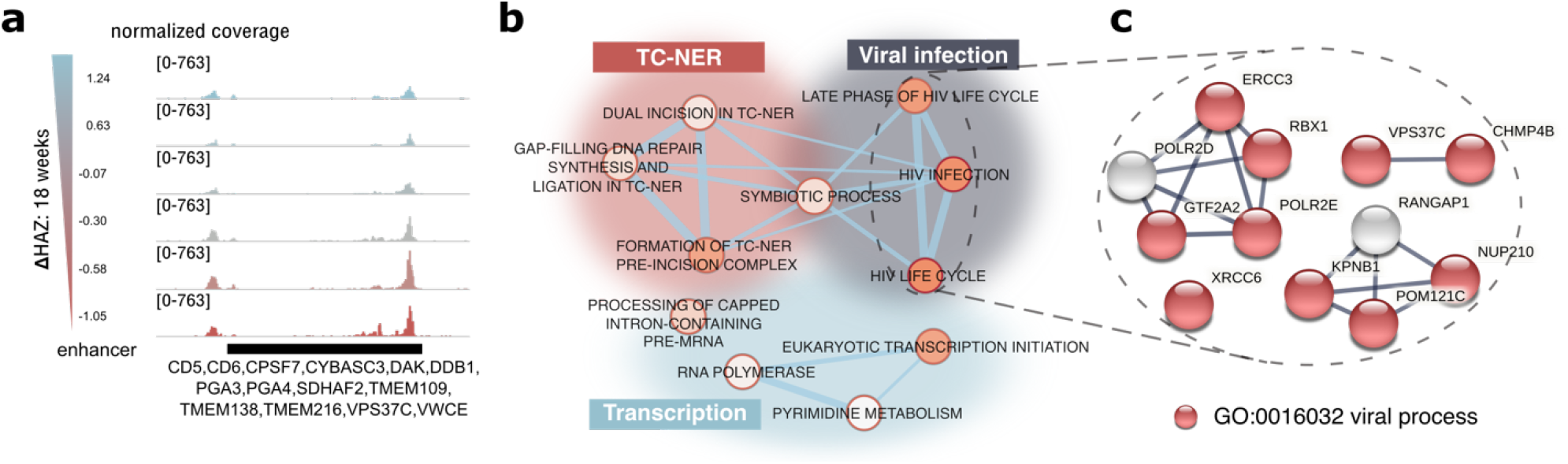
Functional annotation of genes associated with significantly upregulated H3K27ac regions in 18-week-old stunted children. (a) Genome browser snapshot of H3K27ac signal tracks showing a representative region in which the normalized coverage increased with decreasing ΔHAZ (18 wk) score, i.e. H3K27ac was higher in stunted children. The color of individual signal tracks corresponds to the ΔHAZ (18 wk) scores. Genes associated with the highlighted region are labeled. (b) Biological terms (databases: GO biological process, KEGG, Rectome, WikiPathways) that were significantly enriched for genes associated with differential H3K27ac regions in 18-week-old children. Each node is a biological term, the node size corresponds to the term size, the node color indicates the significance of enrichment (bright orange: low q-value, light orange: higher q-value, all q-values < 0.5), and the edges are based on the number of genes shared between terms. Groups of terms were manually organized in different colored clusters with the summary term for the cluster shown in the label with the same color. (c) Gene network constructed with STRING (see Methods) using genes found in the highlighted biological terms in (b). Red genes belong to the “viral processes” GO term.

### Functional assessment of differential H3K27ac regions in 18-week-old children

We next extended the exploration of possible cell subtype specificity described above to determine whether the differentially-affected regions were particularly important for a specific cell type. The normalized chromatin accessibility signal from different cell types was mapped onto these regions, as shown in Additional file 2: Fig. S6a. The results in the top panel of Additional file 2: Fig. S6b show enrichment in blood specific open chromatin regions, however, there was no indication of a particular blood subpopulation driving the H3K27ac changes.

Next, we looked for enrichment of histone marks and DNA-binding factors within the significantly affected regions (Additional file 2: Fig. S7a,b, Additional file 6: Table S5, Additional file 7: Table S6). As expected, we saw the highest enrichment in H2K27ac (Additional file 2: Fig. S7a) followed by other activating histone marks. The only highly enriched repressive histone mark was mono-ubiquitinated histone H2A at lysine 119 (H2AK119) (25), which showed up only in stem cells. Interestingly, the mark with the second most significant enrichment was dimethylation of histone H3 on lysine 4 (H3K4me2), which has been previously shown to be associated with cis regulatory regions driving cell-specific gene expression (26), and also with DNA repair (27). This observation is consistent with the conclusion that stunted children have upregulated DNA repair pathways in early infancy. The top enriched DNA-binding factors (Additional file 2: Fig. S7b) mostly play roles in activation of T-cells and B-cells, as well as more general roles in cellular development.

### Gene targets of downregulated enhancer regions in stunting at 1 year of age

To provide insight into epigenetic changes in 1-year-old stunted children, we performed detailed functional analyses of 341 regions with reduced H3K27ac levels in stunted compared to control children at one year of age. These regions were identified because of the positive correlation of H3K27ac with ΔHAZ (1 yr) score. An example of such a region is shown in Fig. 4a along with the list of its gene targets. Functional annotation of the set of target genes is presented in the network in Fig. 4b, in which we were able to identify functional submodules of pathways downregulated in stunting. Compared to the results from 18-week-old children suggesting an increased immune system response in stunting, stunting status at 1-year was associated with downregulated immune system pathways, suggesting possible immune system exhaustion, or “immunoparalysis” due to prolonged infections (6,28), or problems in immune system development, as discussed above (Additional file 2: Fig. S5c). Similar to the immune system submodule, DNA damage repair pathways were predicted to be downregulated in 1-year-old stunted children in contrast to their upregulation in 18-week-old infants destined to become stunted. Submodules identified uniquely in data from 1-year-old children include negative epigenetic regulation (specifically methylation), Rho GTPase signaling, oxidative stress pathways, cellular organization, and metabolic pathways. Apart from general macromolecule metabolic processes, we were able to identify enrichment in one-carbon metabolic pathways, a simplified diagram of which is shown in Fig. 4c with target genes associated with downregulated H3K27ac regulatory regions listed in light blue.

**Fig. 4.**
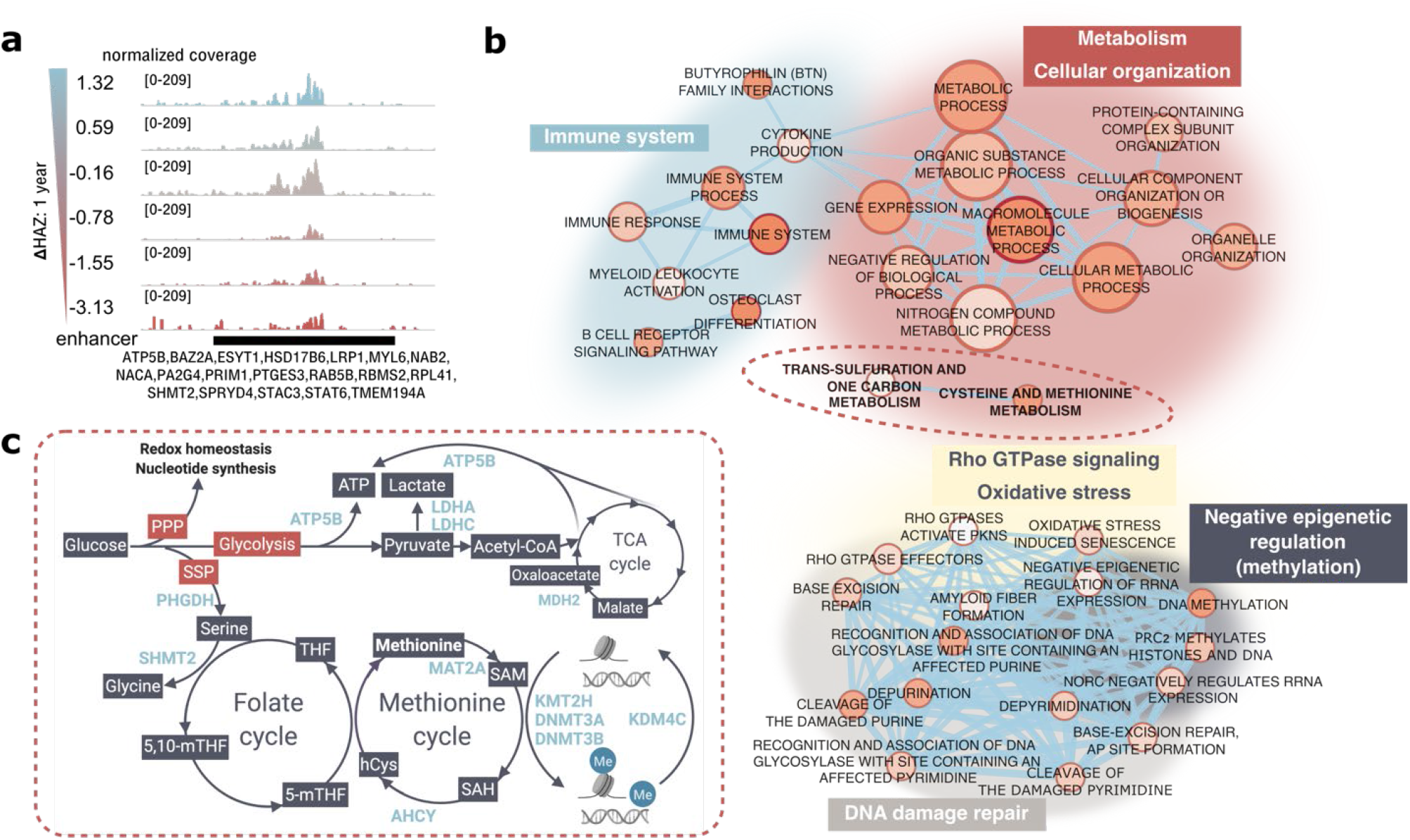
Functional annotation of genes associated with significantly downregulated H3K27ac regions in 1-year-old stunted children. (a) Genome browser screenshot showing a representative region in 1 year old children in which read coverage decreased with descreasing ΔHAZ (1 year) score, i.e. H3K27ac was lower in stunted children. Track colors correspond to the child’s ΔHAZ (1 year) score. Genes associated with the region are labeled. (b) Network of biological terms as in **Fig**. 3b applied on data from 1-year old children. (c) Genes from highlighted biological terms in (b) are shown in a pathway diagram. PPP – pentose phosphate pathway, SSP – serine synthesis pathway.

### Region-centered analysis of downregulated H3K27ac regulatory regions in 1-year-old stunted children

As for the H3K27ac regions identified in 18-week-old children, we wondered if the significantly downregulated regions in 1-year-old children might be enriched in open chromatin regions of a specific cell type. The data shown in the bottom panel of Additional file 2: Fig. S6b indicates that these regions may be particularly important in CD14^+^ monocytes and CD1c^+^ dendritic cells.

Enrichment analysis for histone marks (Additional file 2: Fig. S7c, Additional file 8: Table S7) showed again that as expected, H3K27ac was a top hit, followed by mostly activating histone marks with the highest significance in monocytes and macrophages, as well as heterochromatin regions marked by trimethylated lysine 27 on histone H3 (H3K27me3) in T-cells and in B-cell precursors. The top hits in DNA-binding factor enrichment analysis (Additional file 2: Fig. S7d, Additional file 9: Table S8) include transcription factors driving immune responses to cytokines, growth factors, and interferons, again with the highest significance in monocytes. Interestingly, the enrichment analysis showed H3K4me2 (Additional file 2: Fig. S7c) and Jumonji Domain Containing 1C (JMJD1C) (Additional file 2: Fig. S7d) to be significantly enriched, both of which are associated with DNA damage response pathways (27,29) and therefore provide further evidence that stunted children have altered responses to DNA damage.

### Integrative analysis of H3K27ac and H3K4me3 profiles in 1-year-old children

We next sought to integrate findings from this study with our previously reported results for H3K4me3 in stunting (17). While there were no significant differences in H3K4me3 in 18-week-old children, the differences in H3K4me3 in stunted compared to control children were striking at one year of age. In contrast to the unidirectional changes in H3K27ac levels reported here (Fig. 2c), H3K4me3 changed in both directions in relationship to ΔHAZ (1 yr) (Additional file 2: Fig. S8a, Additional file 10: Table S9), such that H3K4me3 was redistributed from TSS-proximal sites to distal locations.

First, we took a gene-centric approach in which significantly affected H3K27ac and H3K4me3 regions were associated with target genes and pathway enrichment analysis was then performed on those genes present in both lists (Fig. 5a). We separately analyzed genes whose H3K4me3 and H3K27ac regions were both downregulated in stunting (341 genes in Fig. 5b), and genes whose H3K4me3 regions were upregulated, but H3K27ac downregulated in stunting (252 genes in Fig. 5d). Functional enrichment of genes for which both marks were downregulated is shown in Fig. 5c, and for genes with upregulated H3K4me3 but downregulated H3K27ac in Fig. 5e. The results in Fig. 5c show predominantly biological terms connected to metabolism and cellular organization, and thus suggest overall reduced metabolic capacity and possible metabolic rewiring in stunted children. Interestingly, one-carbon metabolism was again detected, providing further support for the hypothesis that stunted children suffer from one carbon metabolite deficiency. The list from Fig. 5b contains the LDL receptor related protein 1 (LRP1) gene, whose role in stunting was previously described by us and supported in a mouse model (17). The results in Fig. 5e show terms associated with viral infection, which may suggest that the loss of H3K27ac is at least partially compensated for by gain of H3K4me3 to maintain immunological responses at the transcriptional level to some extent.

**Fig. 5.**
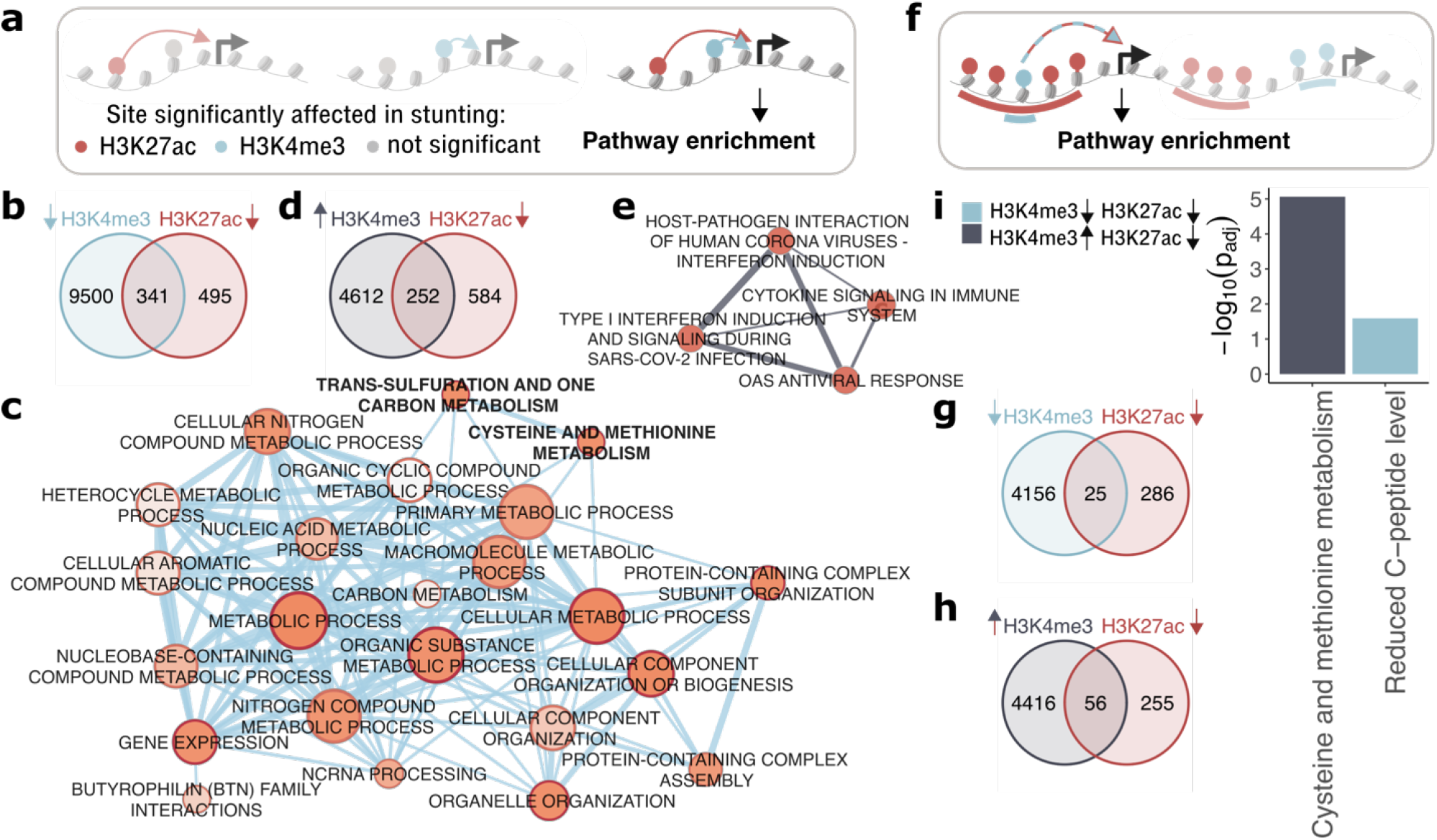
Integrative analysis of H3K27ac and H3K4me3 in 1-year old stunted children. (a) Overview of the “gene-centric” approach in which each differential H3K27ac and H3K4me3 region was assigned to its target gene(s). Only genes predicted to be regulated by both H3K27ac and H3K4me3 were functionally annotated. (b, d) Venn diagrams show overlap of genes regulated by H3K27ac regions that were downregulated in stunting (red circle), and H3K4me3 regions that were downregulated (blue circle) and upregulated (grey circle) in stunting respectively. (c, e) Networks of biological terms as in **Fig**. 3b generated for overlapping genes from (b), and (d) respectively. The color code of network edges corresponds to the color code of H3K4me3 circles in the associated Venn diagrams. (f) Overview of the “region-centric” approach. Significantly affected H3K27ac regions that overlap significantly affected H3K4me3 regions were assigned to target genes, which were functionally annotated. (g, h) Venn diagrams showing intersections of significantly downregulated H3K27ac regions (red circle) in stunting with significantly downregulated (blue circle) or upregulated (grey circle) H3K4me3 regions in stunting. (i) Pathway enrichment of genes associated with overlapping regions from (g, h). Bars are color-coded accordingly with H3K4me3 circles in Venn diagrams.

Next we investigated the relationship between H3K27ac and H3K4me3 at one year of age using a region-centric approach in which significantly affected H3K27ac regions that overlap with significantly affected H3K4me3 regions were associated with genes and these gene sets were investigated for functional enrichment (Fig. 5f). Overall, there was no relationship between the log2-fold changes in H3K27ac and H3K4me3 overlapping regions (Additional file 2: Fig. S8b,c). The concordant overlapping regions that were both decreased in stunting were associated with 25 genes (Fig. 5g), and the discordant regions where stunted children lost H3K27ac but gained H3K4me3 were associated with 56 genes (Fig. 5h). The functional enrichment results for both gene sets are shown in Fig. 5i. The concordant regions were enriched in the C-peptide level pathway. This pathway is associated with low insulin production and is an indicator of disruptions in glucose metabolism leading to diabetes, a disease that tends to emerge in stunted individuals (8,9) (Fig. 1). Discordant regions were found to be enriched in cysteine and methionine metabolism pathways. Importantly, the pattern of H3K4me3 redistribution observed in stunting is similar to changes that result from methionine restriction (17) and both are associated with non-random effects on specific metabolic genes. Collectively, these results provide yet another argument in support of the conclusion that stunted children are deficient in one-carbon metabolism.

Lastly, we restricted the functional annotation to strictly overlapping H3K27ac and H3K4me3 regions (rather than annotating whole H3K27ac regions that overlap with H3K4me3, Additional file 2: Fig. S8d). The pathway enrichment analysis results of the associated genes are presented in Additional file 2: Fig. S8e. Here we observed the association of downregulated H3K4me3 and H3K27ac regions with NOTCH signaling genes, a pathway crucial in development, whereas H3K4me3 regions upregulated within H3K27ac regions show association with genes involved in the acute inflammatory response as well as vitamin B12 metabolism, a component of one-carbon metabolism with a previously reported association with stunting (30).

## Discussion

The results presented here and in our previous work are summarized in the model in Fig. 6. While we did not observe any significant changes in the H3K4me3 profiles of 18-week-old children in association with the emergence of stunting, increased levels of H3K27ac were functionally linked to activation of stress and immune response pathways. Importantly, this occurred before the overt appearance of the stunted phenotype in these children. It has been reported previously that H3K27ac is one of the most dynamic histone marks during immune cell reprogramming (31,32), which could contribute to why changes were observed in H3K27ac but not H3K4me3. Taken together, we suggest that differential enhancer activation in early infancy represents a response to initial encounters with a poor environment, including an increased pathogen load and nutritional challenges.

**Fig. 6.**
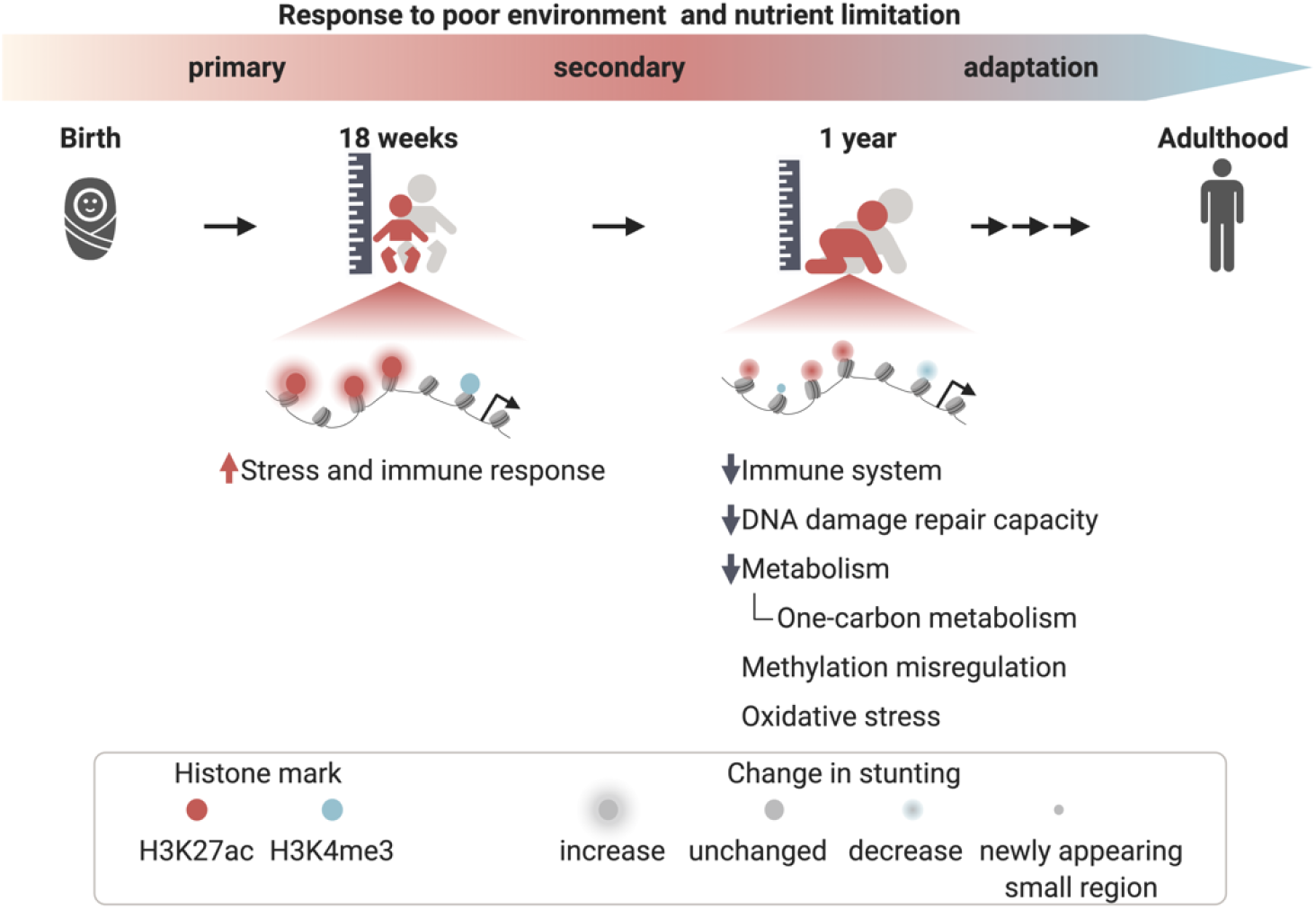
Working model depicting changes to the epigenetic landscape in stunting. The model highlights changes to H3K27ac and H3K4me3 landscapes in 18-week-old and 1-year-old children as they become more severely stunted, along with biological terms associated with these changes.

It is metabolically expensive to maintain immune responses under prolonged periods of increased pathogen exposure (33), and likely unsustainable for growing infants, especially under conditions of nutrient limitation (34). There is a strong possibility that this is one of the reasons for the epigenetic changes we observed in 1-year-old stunted children, which we hypothesize occur as secondary responses to a poor environment and nutrient limitation (Fig. 6). These secondary epigenetic responses consist of global H3K27ac downregulation and a pattern in which H3K4me3 was redistributed from its canonical sites close to TSSs to distal ectopic loci. Functional annotation of both marks indicates that the immune system of stunted children is compromised, which has been well established on a functional level (6,35). The directionality of the epigenetic changes further suggest that immune cells of stunted children become functionally inactive, i.e. exhausted or immune-paralyzed (6,28). Immune system dysfunction is tightly linked to DNA damage and to alterations of DNA damage pathways (36) which are responses to elevated reactive oxygen species (ROS), and oxidative stress (37), all of which appear to be affected in 1-year-old stunted children according to our results (Fig. 4b). Furthermore, aberrant DNA-repair and associated pathways were linked with multiple deficiencies including growth retardation and cognitive impairment (22), which are features of the stunted phenotype.

One of our long-term goals is to identify nutritional interventions that address the specific needs of this stunted child population. The changes we detected in H3K27ac in 1-year-old children suggest downregulation of one-carbon metabolic pathways (Fig. 4b,c). This is particularly interesting since our previously published H3K4me3 results uncovered a similarity between the pattern in stunted children and the pattern in methionine-starved cells (38). We further provide evidence through H3K27ac-H3K4me3 integrative analyses that a deficiency in one-carbon metabolism plays an important role in stunting, as both marks are altered at genes in this pathway in stunted children (Fig. 5c,i). Methionine is an essential amino acid and a precursor for generating the methyl group donor, SAM. High quality protein is metabolically expensive, and a plausible hypothesis is that stunted children are deficient in one carbon metabolites as a result of deficient methionine consumption. Such a deficiency could be addressed through increased availability of complete protein or methionine supplementation in the diet. The consequences of reduced methionine levels on DNA methylation have been previously discussed in acute undernutrition (39). The roles of one-carbon metabolite levels on DNA methylation during pregnancy have also been studied, although many questions remain (40). However, the epigenetic impact of chronic one-carbon metabolite insufficiency in early infancy as we propose here has not been explored. Intriguingly, most of the genes highlighted in Fig. 4c, which are associated with significant loss of H3K27ac in stunting were also identified in the list of genes with loss of H3K4me3 in stunting. The distribution of the affected genes within the pathway (Fig. 4c) may suggest i) methionine deficiency as discussed above, ii) changes in glycolytic flux, and with it associated iii) impairment in serine production. Alterations in glycolysis and serine biosynthesis have been associated with cognitive impairment (41), a common health issue in stunting. Furthermore ii) and iii) have been previously linked with some of the modules shown in Fig. 4b. For example, serine levels have been shown to impact senescence (note the oxidative stress-induced senescence term in Fig. 4b) (42), and enzymes in the glycolytic pathway such as lactate dehydrogenase A (LDHA) have been shown to be important in immune cell activation (43). Intriguingly, the biological responses associated with these metabolites and those that are also observed in stunted children have been shown to be at least partially reversed by metabolite supplementation (38,42,44).

Although our ChIP-seq data were obtained from PBMCs, we see little evidence that the overall pattern changes reported here are attributable to changes in a particular cell subtype within these samples. The data were analyzed in two complementary ways: i) we mapped the normalized ChIP-seq signal from individual samples onto cell type-specific enhancers defined by Andersson et al. (2014) (21) (Additional file 2: Fig. S4), and ii) we mapped the normalized cell type-specific chromatin accessibility signals onto significantly affected regulatory elements (Additional file 2: Fig. S6). The results indicate that these changes are not cell type-specific, a conclusion supported by our prior results (17). A possible exception is shown in Additional file 2: Fig. S6b, which indicates that differentially affected H3K27ac regions in one year old children show slightly higher enrichment in open chromatin regions of monocytes and dendritic cells compared to other subpopulations. These findings are consistent with results from a cohort of stunted infants in Zimbabwe, which showed a decline of the CD14^+^ monocyte population in 1-year-old stunted children (45), as well as results from an animal model of protein malnutrition in which protein-deficiency led to dysfunctional dendritic cells and non-responsiveness to vaccination (44).

## Conclusions

We analyzed H3K27ac, a histone mark associated with active enhancers in PBMCs of healthy and stunted infants aged 18 weeks and 1 year. We identified sites with increased H3K27ac levels in 18-week-old children destined to become stunted, and conversely decreased H3K27ac levels in 1-year-old children who became stunted. Functional analyses associated these sites primarily with immune system and stress response genes. In addition, the data from 1-year-old children point to general changes in metabolic capacity and specifically suggest alterations in one-carbon metabolism that we hypothesize may result from nutritional insufficiency in one carbon precursors.

## Methods

### Human peripheral blood mononuclear cells (PBMC) samples

Deidentified PBMC samples from 18-week-old and 1-year-old children enrolled in the PROVIDE study (19) were obtained in collaboration with icddr,b (International Centre for Diarrhoeal Disease Research, Bangladesh) in Dhaka, Bangladesh. The study was approved by the Ethical Review Board of icddr,b (FWA 00001468) and the Institutional Review Boards of the University of Virginia (FWA 00006183) and the University of Vermont (FWA 00000727). Within 7 d after giving birth, screening for eligibility and study consenting occurred in the household by trained Field Research Assistants. Informed consent was obtained for all participating children (trial registration: ClinicalTrials.gov NCT01375647).

### H3K27ac ChIP-seq

ChIP-seq experiments were performed as previously described in detail in Uchiyama et al. (2018) (17). In brief, PBMC samples from 18-week-old and 1-year-old children were fixed with formaldehyde, chromatin isolated and sheared, 0.02% (microgram/microgram) *Drosophila* chromatin (cat #53083, Active Motif, Carlsbad, CA, USA) was added for spike-in normalization (46). DNA fragments were isolated by immunoprecipitation with H3K27ac antibodies (12.5 μl per 100 μg chromatin protein solution, cat# C15410-196, lot# A1723-0041D, Diagenode, Denville, NJ, USA), and sequencing libraries constructed using the Illumina TruSeq ChIP Library Preparation Kit following the manufacturer instructions. Libraries were sequenced using an Illumina NextSeq500 instrument with high capacity cartridge in the University of Virginia DNA Sciences Core Facility, yielding between 41 million to 72 million 150 bp single-end reads per sample.

### ChIP-seq data preprocessing

Datasets were generated from samples from 18-week-old (14 samples) and 1-year old (22 samples) children, including both males and females and with a broad spectrum of HAZ and ΔHAZ scores (Fig. 2a, Additional file 2: Fig. S1a,j). Raw H3K27ac sequences were aligned to the human genome (version hg19) with bowtie2 (2.2.6.) (47) using default settings. The resulting files were processed to remove unmapped reads and then converted to BAM format using Samtools (0.1.19-4428cd) (48). ENCODE-defined blacklisted sites (49) were removed using bedtools (v2.26.0) *intersect* (50). Quality of each dataset was assessed with FastQC (v0.11.5) (51) in combination with MultiQC (1.2) (52). Peaks identifying H3K27ac enriched regions were called using MACS2 (2.1.1.20160309) (53) against input with parameters set to --broad --broad-cutoff 0.01. Separately identified peaks with MACS2 settings --broad --broad-cutoff 0.05 were stitched together into putative enhancers and superenhancers with ROSE (54,55) with recommended parameters -s 12500 -t 2500. Based on visual inspection of peaks called only with MACS2 and of enhancers called with ROSE in Integrated Genomics Viewer (IGV) (56), enhancers called with ROSE were used for construction of count tables used in downstream differential analysis. To create the count table BED files with enhancer coordinates were merged into a single BED file using bedtools *merge* with default parameters, this was done for the two age groups separately, providing separate sets of putative enhancers for 18-week-old and 1-year-old children. Coverage of individual samples over the final region sets was then enumerated with bedtools *multicov* with default settings, providing separate count tables for each age group, which were then used as an input for differential analysis described in further section of methods. Count tables were also created in the same way for superenhancers.

Sequencing reads were also mapped to the *Drosophila* dm6 genome to obtain read counts for spike-in normalization factors. All necessary hg19 and dm6 files including genome indexes, chromosome sizes, etc. were obtained with refgenie (0.9.3) (57).

To visually inspect individual datasets and create genome browser snapshots, normalized bigWig files were created from BAM files using bedtoos *genomcov* with argument -scale set to a normalization factor for a given sample (see section on data normalization below about calculation of normalization factors) followed by use wigToBigWig tool (58).

### Data normalization

Prior to differential analysis, it is essential to correctly normalize count tables. *Drosophila* spike-in chromatin was used in our experimental set up to correct for global changes (Additional file 2: Fig. S2a). If the ratio of human to *Drosophila* chromatin stays the same, then the percentage of reads mapped to *Drosophila* genome out of all sequencing reads, should be different only if there is a global gain or loss of a histone mark under certain conditions (e.g. in stunted children), as showed in illustration in Additional file 2: Fig. S2b,c. In case that there is no global change between conditions, the percentage of recovered *Drosophila* sequences should be roughly the same (Additional file 2: Fig. S2c). The percentage of reads mapped to *Drosophila* should not be, however, dependent on any other factor, like sequencing depth, which is something that was observed in the data presented here (Additional file 2: Fig. S2d). To eliminate this dependence between sequencing reads and percentage of reads mapped to *Drosophila* out of normalization factors, we regressed out the linear relationship between these two variables and recalculated the normalization factors based on residuals recovered from the linear model. The corrected number of reads mapped to *Drosophila* was then calculated as:

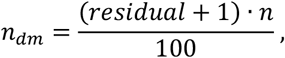

where *n*_*dn*_ is the corrected number of reads mapped to *Drosophila, residual* is a linear model residual corrected by adding 1 to eliminate negative numbers, and *n* is the total number of sequencing reads for a given sample. The final normalization factors were then obtained by dividing n_dm_ with an arbitrary constant, here the constant was chosen to be 500,000. The final values (Additional file 3: Table S2) were used for count table normalization and bigWig file scaling.

### Differential analysis

Differential analysis using normalized count tables was carried out by DESeq2 (1.30.0) (59) with design set to sex + ΔHAZ (18 wk) for 18-week-old children and sex + ΔHAZ (1 yr) for 1-year-old children. This allowed for identification of regions for which the coverage changed dynamically with increasing ΔHAZ score, while controlling for sex differences. Significantly affected regions were then defined as those with an FDR-corrected p-value < 0.05.

The significance of the apparent global shifts in acetylation in stunted children was assessed by performing one-sample two-sided Student’s t-test on log_2_(fold-changes) with μ = 0. Results with p < 0.05 were considered significant.

The alluvial plot assessing changes that occurred in stunted children between 18 weeks and 52 weeks was created by first finding overlaps between regions from the two count tables (18 weeks, 1 year) with bedtools *intersect* function with argument -wao, which identifies overlapping regions, but instead of returning their intersection, the overlapping regions are returned in their original form. This allows connecting results from differential analysis to the regions. Based on log_2_(fold-changes), regions were classified as “up” or “down” in stunting for each group or “not present” if a region in one age group does not overlap with any of the regions defined in the other age group. Data in this form was then plotted as an alluvial plot with ggalluvial (0.12.2) R package (60). Log_2_(fold-changes) connected to overlapping regions (not regions present only in one age group) were also plotted as a scatter plot and their correlation was assessed by calculating Pearson’s correlation coefficient. Regions upregulated at 18-weeks and downregulated at 1-year in stunting selected for functional annotation were selected based on the weighted difference in log2(fold-change) at a given age weighted by FDR-corrected p-values:

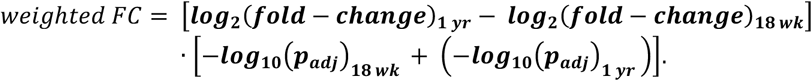

Intersections of the top 10% of these enhancers were functionally annotated as described below.

Principal component analysis (PCA) was carried out as described in the DESeq2 vignette using regularized logarithm transformation; all regions were used for analysis.

### Gene annotation and functional annotation

H3K27ac regions of interest were associated with genes using the EnhancerAtlas 2.0 database (61). A database for PBMCs was created by downloading EnahncerAtlas2 defined enhancer-gene interactions for PBMC-relevant cell types, namely CD4+, CD8+, CD14+, CD19+, CD20+, GM10847, GM12878, GM12891, GM12892, GM18505, GM18526, GM18951, GM19099, GM19193, GM19238, GM19239, GM19240, PBMC cells. Individual text files were reformatted into tab delimited files, in which the first three columns consisted of the region coordinates (chr, start, end) followed by columns with gene identifiers, and cell type. The reformatted files were then concatenated and sorted, giving rise to the final custom PBMC EnhancerAtlas 2.0 database. Regions of interest were assigned to genes by finding overlaps with the regions in the created database.

Functional enrichment analysis using gene lists of interest was performed as recommended by Reimand et al. (2019) (62). Specifically, gene lists were functionally annotated by g:Profiler (63) with data sources restricted to GO biological processes, KEGG, Reactome, and WikiPathways, otherwise with default settings. Outputs from g:Profiler were visually represented in two ways: 1) shorter lists (number of terms ≤ 10) were visualized based on information in CSV files as bar graphs with bar height set to -log_10_(p_adj_), 2) longer lists (number of terms > 10) were visualized as networks using EnrichmentMap (3.3) (64) Cytoscape (3.8.0) (65) application. EnrichmentMap requires the output from g:Profiler in the form of a GEM file along with a GMT file providing information about which gene belongs to a given biological term. GMT files for GO biological processes, Reactome, and WikiPathways were obtained from g:Profiler, and combined with GMT file for KEGG, that was obtained separately from EnrichmentMap website (https://enrichmentmap.readthedocs.io/en/latest/GeneSets.html). The nodes in resulting networks represent individual biological terms, node size correspond to the term size, node color to the significance of the term, and edges were formed based on the number of overlapping genes between the two connected terms. Biological terms were then manually divided into clusters based on their similarity. Due to the large complexity of results obtained in functional annotation of genes associated with regions that changed between 18 weeks and 1 year in stunting, GO biological processes were eliminated in this analysis. Genes involved in the response to viral infection in data from 18-week-old children were validated for functional enrichment with STRING (66) with text mining off.

Enrichment analysis was also performed for regions of interest with LOLA (67) using CISTROME (68,69) databases of transcription factors (termed here as DNA-binding factors) and histone marks filtered for blood cells. The CISTROME database was available only for the hg38 genome assembly, so H3K27ac regions of interest were first converted to hg38 annotation with liftOver (70). Significantly affected regions defined by DESeq2 were used as “userSet”, and all regions from a count table as “userUniverse.” Significant results were identified as those with q-value < 0.5, and presented in form of a dot plot where the x-axis represents different cell types, the y-axis represents different histone marks or DNA-binding factors, and the size along with the color of a dot reflects the most significant q-value for a given cell type – histone mark or cell type -DNA-binding factor combination. Due to the very long list of significant DNA-binding factors, only the top quartile of the results ranked by q-value was visualized.

### Cell-type specificity analysis

Two approaches were employed to identify cell-specific responses from the batch ChIP-seq experiments. In the first approach, average normalized H3K27ac profiles were generated over enhancer regions specific for a given cell type (these include B-cells, T-cells, dendritic cells, macrophages, monocytes, and natural killer cells) defined by Andersson et al. (2014) (21), as shown in Additional file 2: Fig. S4a. To use truly cell specific regions, any intersecting regions among the different cell types were excluded from the analysis. Average profiles were obtained by mapping normalized bigWig files onto the cell specific BED files with deepTools (3.3.1) (71) *computeMatrix* function with arguments –referencePoint center -b 10000 -a 1000 followed by *plotProfile* function. The average profiles were plotted in R with ggplot2 (3.3.2) with smoothing parameters set to geom_smooth(span=0.2).

The GenomicDistributions package (0.99.4) (72) was used as a complementary approach, where normalized open chromatin signal from ENCODE DNase-seq and ATAC-seq experiments across different cell types was mapped onto the H3K27ac regions of interest (Additional file 2: Fig. S6), i.e. onto significant regions identified with DESeq2.

### H3K4me3 data

H3K4me3 dataset from 1-year-old children from Uchiyama et al. (2018) (17) was pre-processed to generate count tables as described in detail in Uchiyama et al. (2018) (17). Differential analysis was performed by DESeq2 (1.30.0) with default normalization and design set to sex + ΔHAZ (1 yr). Significant peaks that were down in stunted children were previously shown to be closer to transcription start sites (TSSs), and were therefore associated with genes by using GREAT (73), while significant peaks that are upregulated in stunted children tended to be further from TSSs and enriched in enhancer regions (17), and were therefore associated with genes by EnhacerAtlas2 -like H3K27ac regions. Venn diagrams for co-regulated genes by H3K4me3 and H3K27ac or for the intersections of the regions were created with VennDiagram package (1.6.20) (74). Pathway enrichment analysis of coregulated genes was performed as described for H3K27ac datasets. Correlation of log2(fold-changes) of intersecting H3K4me3 and H3K27ac regions was evaluated by calculating Pearson’s correlation coefficient.

Illustrations were generated with BioRender (75). Other R packages used in the analysis for data wrangling and plot editing: tidyverse (1.3.0) (76), GenomicRanges (1.42.0) (77), Hmisc (4.4-2) (78), ggpubr (0.4.0) (79), ggmosaic (0.2.0) (80), ggrastr (0.2.1) (81).

## Data Availability

The datasets supporting the conclusions of this article are available in dbGaP repository, phs001073.v2.p1, https://www.ncbi.nlm.nih.gov/gap/. DbGaP identifiers are listed in Additional file 1: Table S1. Important scripts associated with this manuscript are available from: https://github.com/AubleLab/H3K27ac_in_stunting:
(DOI: https://zenodo.org/badge/latestdoi/339230344).

https://www.ncbi.nlm.nih.gov/projects/gap/cgi-bin/study.cgi?study_id=phs001073.v2.p1

## Abbreviations

ChIP-seq: chromatin immunoprecipitation followed by sequencing
CVD: cardiovascular diseases.
FDR: false discovery rate
H2AK119: histone H2A mono-ubiquitination on lysine 119
H3K4me2: histone H3 dimethylation on lysine 4
H3K4me3: histone H3 trimethylation on lysine 4
H3K27ac: histone H3 acetylation on lysine 27
H3K27me3: histone H3 trimethylation on lysine 27
HAZ: height-for-age z-score
ΔHAZ (18 wk): difference in height-for-age z-scores between birth and 18 weeks of age
ΔHAZ (1 yr): difference in height-for-age z-scores between birth and 1 year of age
JMJD1C: Jumonji domain containing 1C
LDHA: lactate dehydrogenase A
LRP1: LDL receptor related protein 1
PBMC: peripheral blood mononuclear cells
PCA: principal component analysis
PPP: pentose phosphate pathway
PROVIDE: performance of rotavirus and oral polio vaccines in developing countries
SAM: S-adenosyl methionine
SSP: serine synthesis pathway
T2D: type 2 diabetes
TC-NER: transcription coupled nucleotide excision repair
TSS: transcription start site
WHO: World Health Organization

## Declarations

### Ethics approval and consent to participate

The study was approved by the Ethical Review Board of icddr,b (FWA 00001468) and the Institutional Review Boards of the University of Virginia (FWA 00006183) and the University of Vermont (FWA 00000727). All samples were analyzed in a de-identified fashion. Informed consent was obtained for all participants (trial registration: ClinicalTrials.gov NCT01375647).

### Consent for publication

Not applicable.

### Availability of data and materials

The datasets supporting the conclusions of this article are available in dbGaP repository, phs001073.v2.p1, https://www.ncbi.nlm.nih.gov/gap/. DbGaP identifiers are listed in Additional file 1: Table S1. Important scripts associated with this manuscript are available from: https://github.com/AubleLab/H3K27ac_in_stunting: (DOI: https://zenodo.org/badge/latestdoi/339230344).

## Authors’ contributions

DA conceived of the study and supervised the analysis; SJS isolated chromatin and prepared libraries for sequencing; KK conducted the analyses, RH and WAP, Jr, conceived of and established the PROVIDE Study; all authors wrote the manuscript.

## Acknowledgements

We are grateful to the mothers and children who participated in the PROVIDE Study. We are also grateful to Stefan Bekiranov for discussions about data normalization, to Nathan Sheffield for discussions and guidance with cell-specific analyses and to Billy Nash for proofreading the manuscript and providing insightful comments.

## List of additional files

**Additional file 1**. Table S1. List of samples and associated metadata.

**Additional file 2**. Supplementary figures (S1-S8).

**Additional file 3**. Table S2. Pre-processing results.

**Additional file 4**. Table S3. H3K27ac differential analysis results for 18-week-old children.

**Additional file 5**. Table S4. H3K27ac differential analysis results for 1-year-old children.

**Additional file 6**. Table S5. Significant histone marks identified with LOLA for 18-week-old children.

**Additional file 7**. Table S6. Significant DNA-binding factors identified with LOLA for 18-week-old children.

**Additional file 8**. Table S7. Significant histone marks identified with LOLA for 1-year-old children.

**Additional file 9**. Table S8. Significant DNA-binding factors identified with LOLA for 1-year-old children.

**Additional file 10**. Table S9. H3K4me3 differential analysis results for 1-year-old children.

## References

1. World Health Organization. Reducing stunting in children: equity considerations for achieving the global targets 2025. Who. 2018.

2. Black RE, Victora CG, Walker SP, Bhutta ZA, Christian P, De Onis M, et al. Maternal and child undernutrition and overweight in low-income and middle-income countries. The Lancet. 2013.

3. Humphrey JH. Child undernutrition, tropical enteropathy, toilets, and handwashing. The Lancet. 2009.

4. Dicker D, Nguyen G, Abate D, Abate KH, Abay SM, Abbafati C, et al. Global, regional, and national age-sex-specific mortality and life expectancy, 1950-2017: A systematic analysis for the Global Burden of Disease Study 2017. Lancet. 2018 Nov 10;392(10159):1684–735.

5. United Nations Children’s Fund (UNICEF), World Health Organization, International Bank for Reconstruction and Development/The World Bank. Levels and trends in child malnutrition: Key Findings of the 2020 Edition of the Joint Child Malnutrition Estimates. 2020th ed. Geneva: World Health Organization; 2020.

6. Bourke CD, Jones KDJ, Prendergast AJ. Current understanding of innate immune cell dysfunction in childhood undernutrition. Vol. 10, Frontiers in Immunology. Frontiers Media S.A.; 2019. p. 1728.

7. Bhutta ZA, Ahmed T, Black RE, Cousens S, Dewey K, Giugliani E, et al. What worksã Interventions for maternal and child undernutrition and survival. Vol. 371, The Lancet. Elsevier B.V.; 2008. p. 417–40.

8. Prendergast AJ, Humphrey JH. The stunting syndrome in developing countries. Paediatr Int Child Health. 2014;

9. Guerrant RL, Deboer MD, Moore SR, Scharf RJ, Lima AAM. The impoverished gut - A triple burden of diarrhoea, stunting and chronic disease. Nat Rev Gastroenterol Hepatol. 2013;10(4):220–9.

10. Grantham-McGregor S, Cheung YB, Cueto S, Glewwe P, Richter L, Strupp B. Developmental potential in the first 5 years for children in developing countries. Lancet. 2007;369(9555):60–70.

11. Guerrant RL, Oriá RB, Moore SR, Oriá MOB, Lima AAAM. Malnutrition as an enteric infectious disease with long-term effects on child development. Nutr Rev. 2008;66(9):487–505.

12. Raman AS, Gehrig JL, Venkatesh S, Chang HW, Hibberd MC, Subramanian S, et al. A sparse covarying unit that describes healthy and impaired human gut microbiota development. Science (80-). 2019;

13. Gehrig JL, Venkatesh S, Chang HW, Hibberd MC, Kung VL, Cheng J, et al. Effects of microbiota-directed foods in gnotobiotic animals and undernourished children. Science (80-). 2019;

14. WHO. Global targets 2025. Glob targets 2025. 2014;

15. Goudet SM, Bogin BA, Madise NJ, Griffiths PL. Nutritional interventions for preventing stunting in children (Birth to 59 months) living in urban slums in low-and middle-income countries (LMIC). Cochrane Database Syst Rev. 2019;

16. Dai Z, Ramesh V, Locasale JW. The evolving metabolic landscape of chromatin biology and epigenetics. Vol. 21, Nature Reviews Genetics. Nature Research; 2020. p. 737–53.

17. Uchiyama R, Kupkova K, Shetty SJ, Linford AS, Pray-Grant MG, Wagar LE, et al. Histone H3 lysine 4 methylation signature associated with human undernutrition. Proc Natl Acad Sci U S A. 2018 Nov 12;115(48):E11264–E11273.

18. Creyghton MP, Cheng AW, Welstead GG, Kooistra T, Carey BW, Steine EJ, et al. Histone H3K27ac separates active from poised enhancers and predicts developmental state. Proc Natl Acad Sci U S A. 2010 Dec 14;107(50):21931–6.

19. Kirkpatrick BD, Colgate ER, Mychaleckyj JC, Haque R, Dickson DM, Carmolli MP, et al. The “Performance of Rotavirus and Oral Polio Vaccines in Developing Countries” (PROVIDE) study: description of methods of an interventional study designed to explore complex biologic problems. Am J Trop Med Hyg. 2015 Apr;92(4):744–51.

20. Paauw ND, Lely AT, Joles JA, Franx A, Nikkels PG, Mokry M, et al. H3K27 acetylation and gene expression analysis reveals differences in placental chromatin activity in fetal growth restriction. Clin Epigenetics. 2018 Jun 26;10(1):85.

21. Andersson R, Gebhard C, Miguel-Escalada I, Hoof I, Bornholdt J, Boyd M, et al. An atlas of active enhancers across human cell types and tissues. Nature. 2014 Mar 26;507(7493):455–61.

22. Lans H, Hoeijmakers JHJ, Vermeulen W, Marteijn JA. The DNA damage response to transcription stress. Vol. 20, Nature Reviews Molecular Cell Biology. Nature Research; 2019. p. 766–84.

23. Guerrant RL, Deboer MD, Moore SR, Scharf RJ, Lima AAM. The impoverished gut - A triple burden of diarrhoea, stunting and chronic disease. Vol. 10, Nature Reviews Gastroenterology and Hepatology. Nature Publishing Group; 2013. p. 220–9.

24. Weitzman MD, Weitzman JB. What’s the damageã The impact of pathogens on pathways that maintain host genome integrity. Vol. 15, Cell Host and Microbe. Cell Press; 2014. p. 283–94.

25. Tamburri S, Lavarone E, Fernández-Pérez D, Conway E, Zanotti M, Manganaro D, et al. Histone H2AK119 Mono-Ubiquitination Is Essential for Polycomb-Mediated Transcriptional Repression. Mol Cell. 2020 Feb 20;77(4):840-856.e5.

26. Pekowska A, Benoukraf T, Ferrier P, Spicuglia S. A unique H3K4me2 profile marks tissue-specific gene regulation. Genome Res. 2010 Nov 1;20(11):1493–502.

27. Wang S, Meyer DH, Schumacher B. H3K4me2 regulates the recovery of protein biosynthesis and homeostasis following DNA damage. Nat Struct Mol Biol. 2020 Dec 1;27(12):1165–77.

28. Wherry EJ, Kurachi M. Molecular and cellular insights into T cell exhaustion. Vol. 15, Nature Reviews Immunology. Nature Publishing Group; 2015. p. 486–99.

29. Watanabe S, Watanabe K, Akimov V, Bartkova J, Blagoev B, Lukas J, et al. JMJD1C demethylates MDC1 to regulate the RNF8 and BRCA1-mediated chromatin response to DNA breaks. Nat Struct Mol Biol. 2013 Dec 17;20(12):1425–33.

30. Hasan MM, Fahim SM, Das S, Gazi MA, Mahfuz M. Association of Plasma Low-Density Lipoprotein Receptor-Related Protein-1 (LRP1) with Undernutrition: A Case-Control Study in Bangladeshi Adults. Res Sq. 2020 Aug 13;

31. Saeed S, Quintin J, Kerstens HHD, Rao NA, Aghajanirefah A, Matarese F, et al. Epigenetic programming of monocyte-to-macrophage differentiation and trained innate immunity. Science (80-). 2014 Sep 26;345(6204).

32. Novakovic B, Habibi E, Wang SY, Arts RJW, Davar R, Megchelenbrink W, et al. β-Glucan Reverses the Epigenetic State of LPS-Induced Immunological Tolerance. Cell. 2016 Nov 17;167(5):1354–1368.e14.

33. Lochmiller RL, Deerenberg C. Trade-offs in evolutionary immunology: just what is the cost of immunityã Oikos. 2000 Jan 1;88(1):87–98.

34. Kedia-Mehta N, Finlay DK. Competition for nutrients and its role in controlling immune responses. Vol. 10, Nature Communications. Nature Publishing Group; 2019. p. 1–8.

35. Bourke CD, Berkley JA, Prendergast AJ. Immune Dysfunction as a Cause and Consequence of Malnutrition. Vol. 37, Trends in Immunology. Elsevier Ltd; 2016. p. 386–98.

36. Nakad R, Schumacher B. DNA damage response and immune defense: Links and mechanisms. Vol. 7, Frontiers in Genetics. Frontiers Media S.A.; 2016. p. 147.

37. Barzilai A, Yamamoto KI. DNA damage responses to oxidative stress. Vol. 3, DNA Repair. DNA Repair (Amst); 2004. p. 1109–15.

38. Mentch SJ, Mehrmohamadi M, Huang L, Liu X, Gupta D, Mattocks D, et al. Histone Methylation Dynamics and Gene Regulation Occur through the Sensing of One-Carbon Metabolism. Cell Metab. 2015 Nov 3;22(5):861–73.

39. Schulze K V., Swaminathan S, Howell S, Jajoo A, Lie NC, Brown O, et al. Edematous severe acute malnutrition is characterized by hypomethylation of DNA. Nat Commun. 2019 Dec 1;10(1):1–13.

40. James P, Sajjadi S, Tomar AS, Saffari A, Fall CHD, Prentice AM, et al. Candidate genes linking maternal nutrient exposure to offspring health via DNA methylation: A review of existing evidence in humans with specific focus on one-carbon metabolism. Int J Epidemiol. 2018 Dec 1;47(6):1910–37.

41. Le Douce J, Maugard M, Veran J, Matos M, Jégo P, Vigneron PA, et al. Impairment of Glycolysis-Derived L-Serine Production in Astrocytes Contributes to Cognitive Deficits in Alzheimer’s Disease. Cell Metab. 2020 Mar 3;31(3):503-517.e8.

42. Enriquez-Hesles E, Smith DL, Maqani N, Wierman MB, Sutcliffe MD, Fine RD, et al. A cell non-autonomous mechanism of yeast chronological aging regulated by caloric restriction and one-carbon metabolism. J Biol Chem. 2020 Nov 26;296:100125.

43. Xu K, Yin N, Peng M, Stamatiades EG, Shyu A, Li P, et al. Glycolysis fuels phosphoinositide 3-kinase signaling to bolster T cell immunity. Science (80-). 2021 Jan 22;371(6527):405–10.

44. Niiya T, Akbar SMF, Yoshida O, Miyake T, Matsuura B, Murakami H, et al. Impaired dendritic cell function resulting from chronic undernutrition disrupts the antigen-specific immune response in mice. J Nutr. 2007 Mar 1;137(3):671–5.

45. Prendergast AJ, Rukobo S, Chasekwa B, Mutasa K, Ntozini R, Mbuya MNN, et al. Stunting Is Characterized by Chronic Inflammation in Zimbabwean Infants. John-Stewart GC, editor. PLoS One. 2014 Feb 18;9(2):e86928.

46. Bonhoure N, Bounova G, Bernasconi D, Praz V, Lammers F, Canella D, et al. Quantifying ChIP-seq data: A spiking method providing an internal reference for sample-to-sample normalization. Genome Res. 2014;24(7):1157–68.

47. Langmead B, Salzberg SL. Fast gapped-read alignment with Bowtie 2. Nat Methods. 2012;9(4):357–9.

48. Li H, Handsaker B, Wysoker A, Fennell T, Ruan J, Homer N, et al. The Sequence Alignment/Map format and SAMtools. Bioinformatics. 2009 Aug;25(16):2078–9.

49. Amemiya HM, Kundaje A, Boyle AP. The ENCODE Blacklist: Identification of Problematic Regions of the Genome. Sci Rep. 2019;9(1):9354.

50. Quinlan AR, Hall IM. BEDTools: a flexible suite of utilities for comparing genomic features. Bioinformatics. 2010 Mar 15;26(6):841–2.

51. Andrews S. FastQC: a quality control tool for high throughput sequence data. [Internet]. 2010. Available from: https://www.bioinformatics.babraham.ac.uk/projects/fastqc/

52. Ewels P, Magnusson M, Lundin S, Käller M. MultiQC: summarize analysis results for multiple tools and samples in a single report. Bioinformatics. 2016 Oct 1;32(19):3047–8.

53. Zhang Y, Liu T, Meyer CA, Eeckhoute J, Johnson DS, Bernstein BE, et al. Model-based Analysis of ChIP-Seq (MACS). Genome Biol. 2008;9(9):R137.

54. Lovén J, Hoke HA, Lin CY, Lau A, Orlando DA, Vakoc CR, et al. Selective inhibition of tumor oncogenes by disruption of super-enhancers. Cell. 2013 Apr 11;153(2):320–34.

55. Whyte WA, Orlando DA, Hnisz D, Abraham BJ, Lin CY, Kagey MH, et al. Master transcription factors and mediator establish super-enhancers at key cell identity genes. Cell. 2013 Apr 11;153(2):307–19.

56. Thorvaldsdottir H, Robinson JT, Mesirov JP. Integrative Genomics Viewer (IGV): high-performance genomics data visualization and exploration. Brief Bioinform. 2013 Mar 1;14(2):178–92.

57. Stolarczyk M, Reuter VP, Smith JP, Magee NE, Sheffield NC. Refgenie: a reference genome resource manager. Gigascience. 2020 Feb 1;9(2).

58. Kent WJ, Zweig AS, Barber G, Hinrichs AS, Karolchik D. BigWig and BigBed: enabling browsing of large distributed datasets. Bioinformatics. 2010 Sep 1;26(17):2204–7.

59. Love MI, Huber W, Anders S. Moderated estimation of fold change and dispersion for RNA-seq data with DESeq2. Genome Biol. 2014 Dec 5;15(12):550.

60. Brunson J. ggalluvial: Layered Grammar for Alluvial Plots. J Open Source Softw. 2020 May 21;5(49):2017.

61. Gao T, Qian J. EnhancerAtlas 2.0: An updated resource with enhancer annotation in 586 tissue/cell types across nine species. Nucleic Acids Res. 2020 Jan 1;48(D1):D58–64.

62. Reimand J, Isserlin R, Voisin V, Kucera M, Tannus-Lopes C, Rostamianfar A, et al. Pathway enrichment analysis and visualization of omics data using g:Profiler, GSEA, Cytoscape and EnrichmentMap. Nat Protoc. 2019 Feb 1;14(2):482–517.

63. Raudvere U, Kolberg L, Kuzmin I, Arak T, Adler P, Peterson H, et al. G:Profiler: A web server for functional enrichment analysis and conversions of gene lists (2019 update). Nucleic Acids Res. 2019 Jul 1;47(W1):W191–8.

64. Merico D, Isserlin R, Stueker O, Emili A, Bader GD. Enrichment Map: A Network-Based Method for Gene-Set Enrichment Visualization and Interpretation. Ravasi T, editor. PLoS One. 2010 Nov 15;5(11):e13984.

65. Shannon P, Markiel A, Ozier O, Baliga NS, Wang JT, Ramage D, et al. Cytoscape: A software Environment for integrated models of biomolecular interaction networks. Genome Res. 2003 Nov;13(11):2498–504.

66. Szklarczyk D, Gable AL, Lyon D, Junge A, Wyder S, Huerta-Cepas J, et al. STRING v11: Protein-protein association networks with increased coverage, supporting functional discovery in genome-wide experimental datasets. Nucleic Acids Res. 2019 Jan 8;47(D1):D607–13.

67. Sheffield NC, Bock C. LOLA: Enrichment analysis for genomic region sets and regulatory elements in R and Bioconductor. Bioinformatics. 2016 Feb 15;32(4):587–9.

68. Mei S, Qin Q, Wu Q, Sun H, Zheng R, Zang C, et al. Cistrome Data Browser: A data portal for ChIP-Seq and chromatin accessibility data in human and mouse. Nucleic Acids Res. 2017 Jan 1;45(D1):D658–62.

69. Zheng R, Wan C, Mei S, Qin Q, Wu Q, Sun H, et al. Cistrome Data Browser: Expanded datasets and new tools for gene regulatory analysis. Nucleic Acids Res. 2019 Jan 8;47(D1):D729–35.

70. Hinrichs AS, Karolchik D, Baertsch R, Barber GP, Bejerano G, Clawson H, et al. The UCSC Genome Browser Database: update 2006. Nucleic Acids Res. 2006;34(Database issue):D590.

71. Ramírez F, Dündar F, Diehl S, Grüning BA, Manke T. DeepTools: A flexible platform for exploring deep-sequencing data. Nucleic Acids Res. 2014 Jul 1;42(W1):W187.

72. Kupkova K, Verdezoto J, Smith JP, Stolarczyk M, Danehy T, Lawson JT, et al. GenomicDistributions: fast analysis of genomic intervals with Bioconductor [Internet]. 2020. Available from: http://code.databio.org/GenomicDistributions

73. McLean CY, Bristor D, Hiller M, Clarke SL, Schaar BT, Lowe CB, et al. GREAT improves functional interpretation of cis-regulatory regions. Nat Biotechnol. 2010 May;28(5):495–501.

74. Chen H, Boutros PC. VennDiagram: A package for the generation of highly-customizable Venn and Euler diagrams in R. BMC Bioinformatics. 2011 Jan 26;12(1):35.

75. BioRender [Internet]. [cited 2021 Feb 18]. Available from: https://biorender.com/

76. Wickham H, Averick M, Bryan J, Chang W, McGowan L, François R, et al. Welcome to the Tidyverse. J Open Source Softw. 2019 Nov 21;4(43):1686.

77. Lawrence M, Huber W, Pagès H, Aboyoun P, Carlson M, Gentleman R, et al. Software for Computing and Annotating Genomic Ranges. Prlic A, editor. PLoS Comput Biol. 2013 Aug 8;9(8):e1003118.

78. Harrell FEJ, Dupont C. Hmisc: Harrell Miscellaneous. 2021.

79. Kassambara A. ggpubr: “ggplot2” Based Publication Ready Plots. 2020.

80. Jeppson H, Hofmann H, Cook D. ggmosaic: Mosaic Plots in the “ggplot2” Framework. R package version 0.2.0. 2021.

81. Petukhov V, van den Brand T, Biederstedt E. ggrastr: Raster Layers for “ggplot2”. R package version 0.2.1. [Internet]. 2021. Available from: https://github.com/VPetukhov/ggrastr

