## Additional file 2 for "Histone H3 lysine 27 acetylation profile undergoes two global shifts in undernourished children and suggests one-carbon metabolite insufficiency"

### Supplementary figures

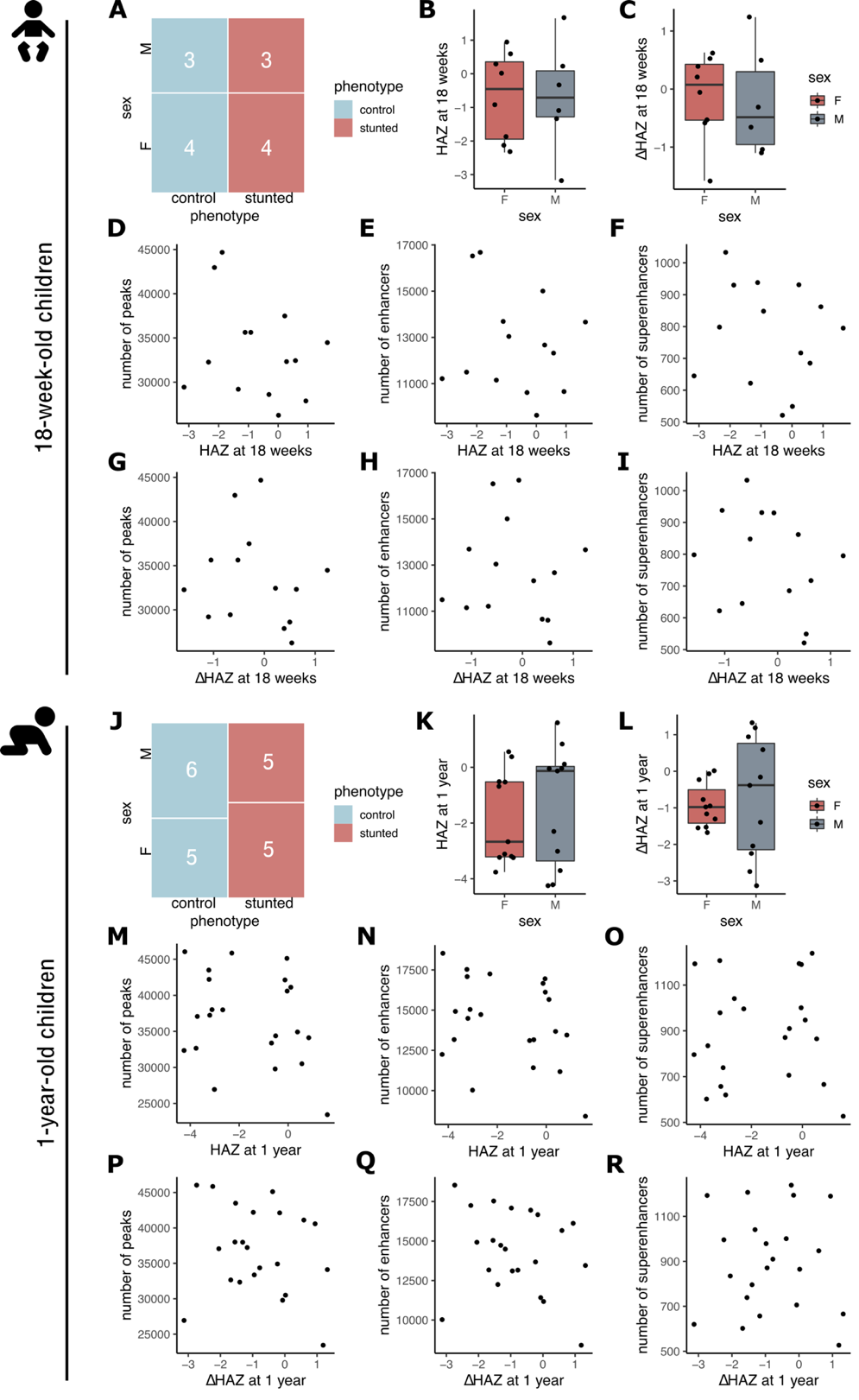

Fig. S1. (A) Mosaic plot showing stunting status (defined by HAZ at 1 year) vs. sex distribution of 18-week-old children whose samples were used in this study. (B, C) Box plots of HAZ (18 wk) and ΔHAZ (18 wk) scores of 18-week-old males and females from this study. (D – F) Scatter plot showing relationship between HAZ (18 wk) and number of identified peaks (D), enhancers (E), and superenhancers (F). (G – I) Scatter plot showing relationship between ΔHAZ (18 wk) and number of identified peaks (G), enhancers (H), and superenhancers (I). (J – R) Same as in (A – I) for 1-year-old children, HAZ, and ΔHAZ scores at 1 year. Boxplots show median, first and third quartile and 95% confidence interval of median.

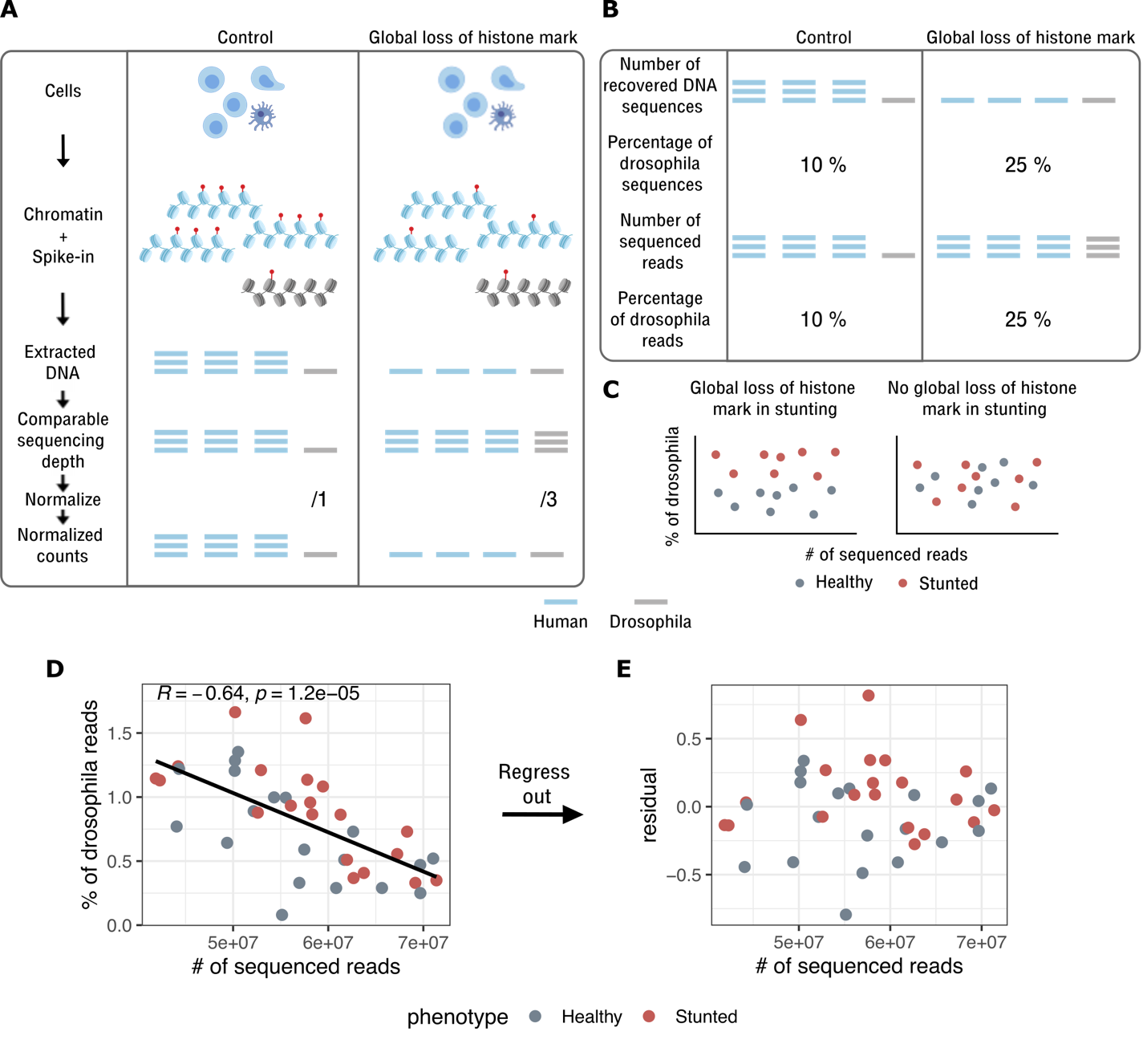

Fig. S2. (A) Motivation for use of spike-in chromatin for ChIP-seq data normalization in order to detect global changes to histone marks. Blue chromatin – primary samples (human), grey chromatin – spike-in (drosophila chromatin). (B) Illustration showing, how sequencing depth does not influence the percentage of spike-in reads. The percentage of spike-in reads is influenced purely by global loss or gain of a histone mark. (C) Hypothetical examples where data from stunted children indicate global loss of a histone mark (left), or no difference in histone mark levels globally across the genome (right). In either case, the percentage of spike-in reads is not correlated with the total number or reads obtained from sequencing. (D) Data from this study showing that the percentage of spike-in reads is dependent on the yield of reads from sequencing. (R indicates Pearson’s correlation coefficient, p is p-value associated with the coefficient). Linear model summary: p = 1.20e-05, multiple R^2^: 0.41, adjusted R^2^: 0.39. (E) Percentage of spike-in reads was corrected by regressing out the linear relationship between the total number of reads and the percentage of spike-in reads (see Methods). Linear model residuals (+1 to eliminate negative numbers) were then used as corrected spike-in percentages.

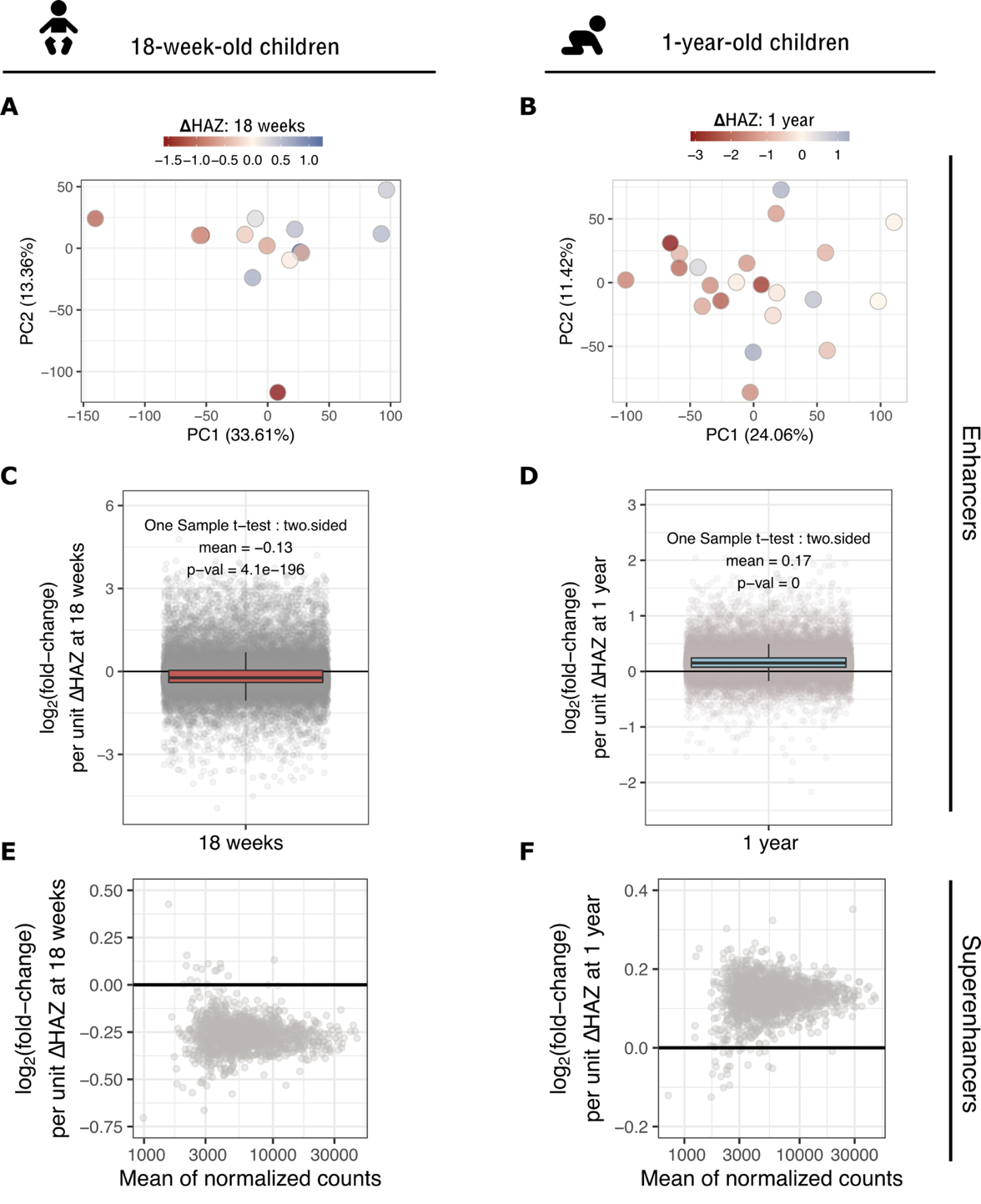

Fig. S3. (A, B) Principal component analysis (PCA) plot of H3K27ac datasets from 18-week-old and 1-year-old children, respectively. PCA was performed on count tables in identified enhancer regions. Each dot represents a child’s sample and is colored based on the ΔHAZ score at a given age. Variance explained by each PC is indicated in parenthesis. (C, D) Box-plots showing, how H3K27ac was globally upregulated in 18-week-old stunted children, and globally downregulated in 1-year-old children, respectively. Each dot is a H3K27ac region with log_2_(fold-change) per unit increase of ΔHAZ score at a given age, i.e. positive log_2_(fold-change) corresponds to a downregulated region in children destined to become stunted and vice versa. Boxes are color-coded in accordance with Fig. 2, where red corresponds to upregulation and blue to downregulation in stunted children. (E, F) MA-plots, as in Fig. 2c,d, but showing changes in identified superenhancer regions in 18-week-old and 1-year-old children. Lack of colored dots indicates that none significant changes were detected. Boxplots show median, first and third quartile and 95% confidence interval of median.

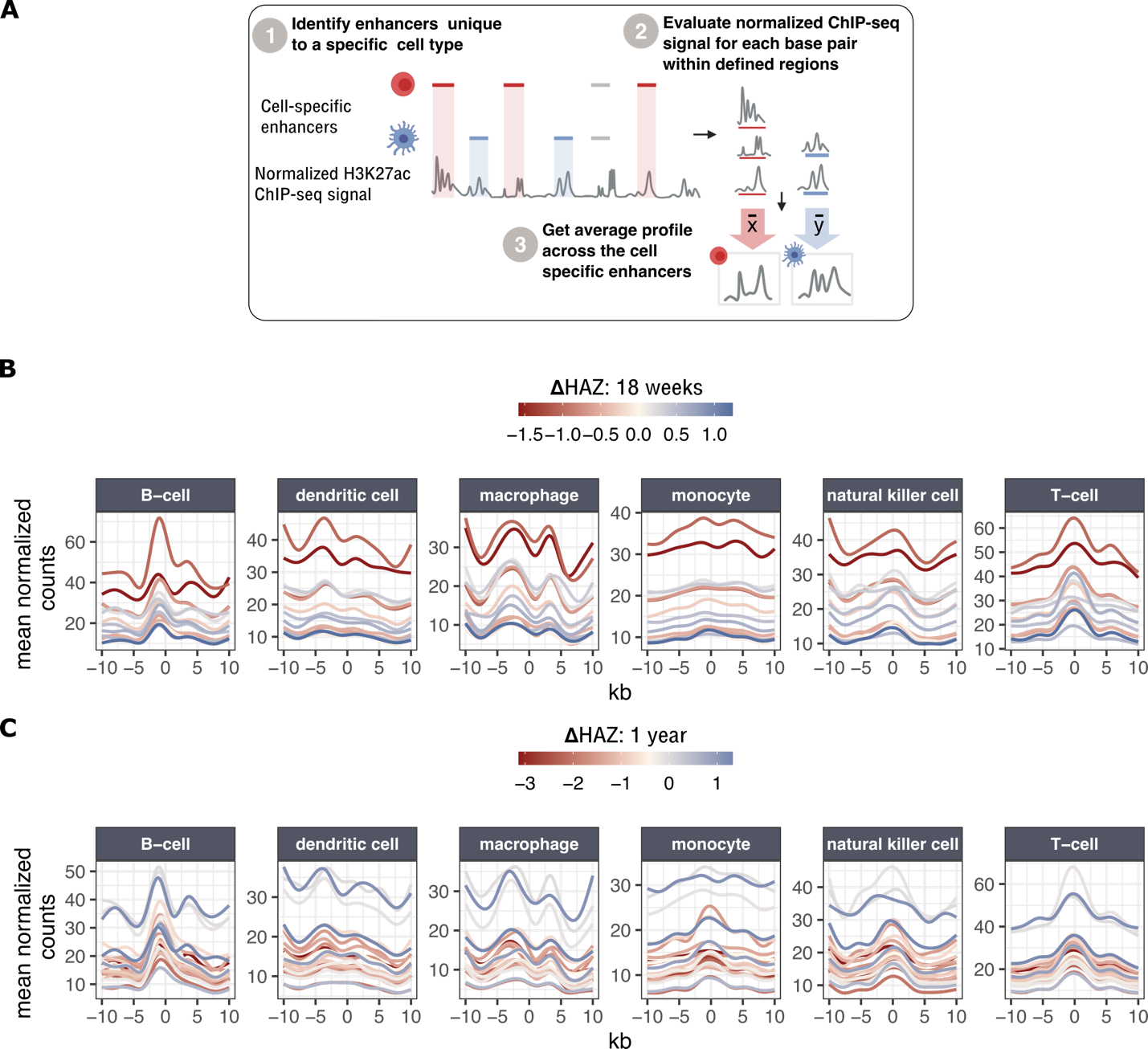

Fig. S4. (A) Workflow for obtaining normalized average ChIP-seq profiles across cell-specific enhancers defined by Andersson et al. (2014) (21). Only unique regions that do not overlap among different cell types were considered in the analysis. (B, C) Normalized average profiles across PBMC-relevant cell types for 18-week-old (B) and 1-year-old children (C). Each line corresponds to a normalized average profile of a child with ΔHAZ score corresponding to the color code in the legend.

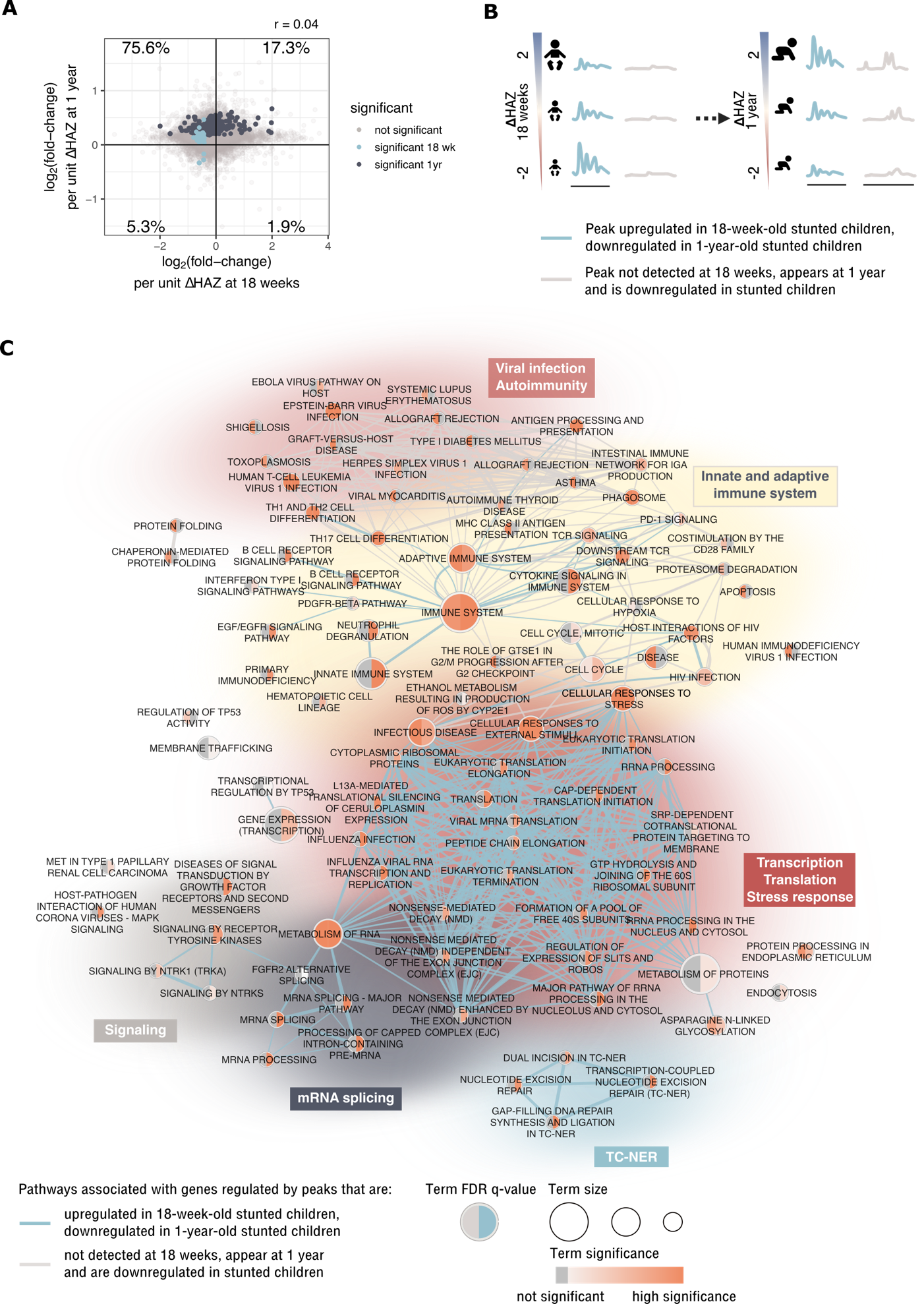

Fig. S5. (A) Scatter plot showing comparison of log_2_(fold-changes) per unit increase of ΔHAZ score at a given age in 1-year-old children vs. 18-week-old children. Compared here are regions that overlap in the two datasets (regions that appeared or disappeared between 18 weeks and 1 year have log_2_(fold-change) value only for 1 age category). Colored are regions that were significantly affected at a given age (none of the regions were found to be significant in both ages). The percentage in each corner indicates the percentage of points (regions) that fall into a give quadrant. Pearson correlation coefficient is indicated on top of the plot. (B) Illustration of the two largest categories of regions in the alluvial plot from Fig. 2d – blue: H3K27ac was upregulated in these regions in stunted children at 18 weeks, then became downregulated in the same regions in stunted children at 1 year, grey: H3K27ac enriched regions were not detected in 18-week old children, and were downregulated in 1-year-old children with higher risk of stunting. (C) Top 10% of regions from the two categories illustrated in (B) were selected based on: i) blue regions - highest value of differences in log_2_(fold-changes) between the two ages weighted by the sum of their associated negative log10(p_adj_); ii) grey regions - lowest padj value at 1 year. The network shows functional annotation of both sets of regions (databases: KEGG, Reactome, WikiPathways). Blue edges show relationships between biological terms associated with blue regions, grey edges with grey regions.

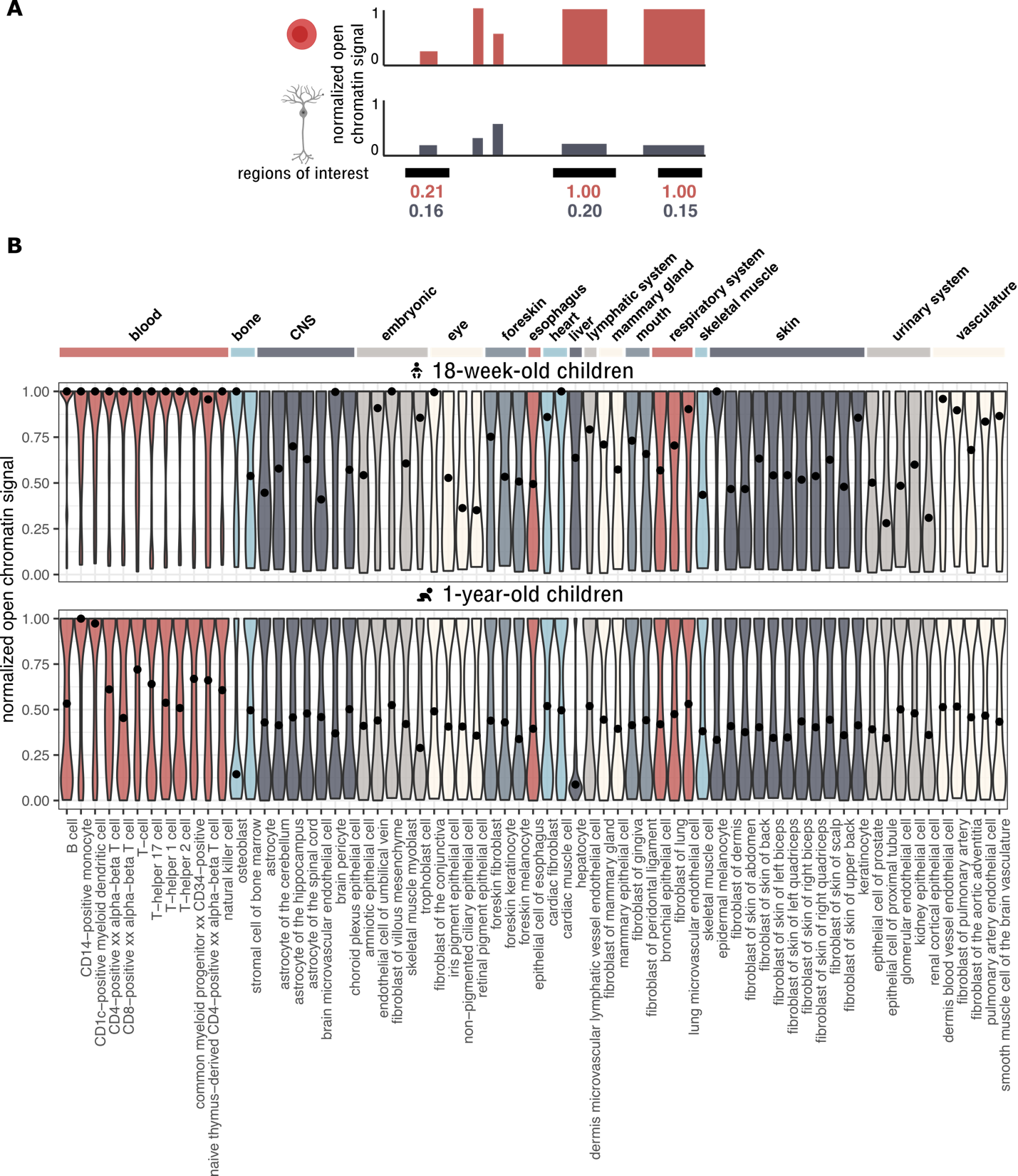

Fig. S6. (A) GenomicDistributions package contains pre-calculated normalized chromatin accessibility values across different cell types in pre-defined regions. These pre-calculated values were projected onto regions of interest. High values for a given cell type indicate that the regions of interest are particularly active in/ important for that cell type, compared to other cell types. (B) Violin plots show distribution of normalized chromatin accessibility values across different cell types in differential regions identified in 18-week-old children (top) or 1-year-old children (bottom). Black dots on top of violin plots represent medians.

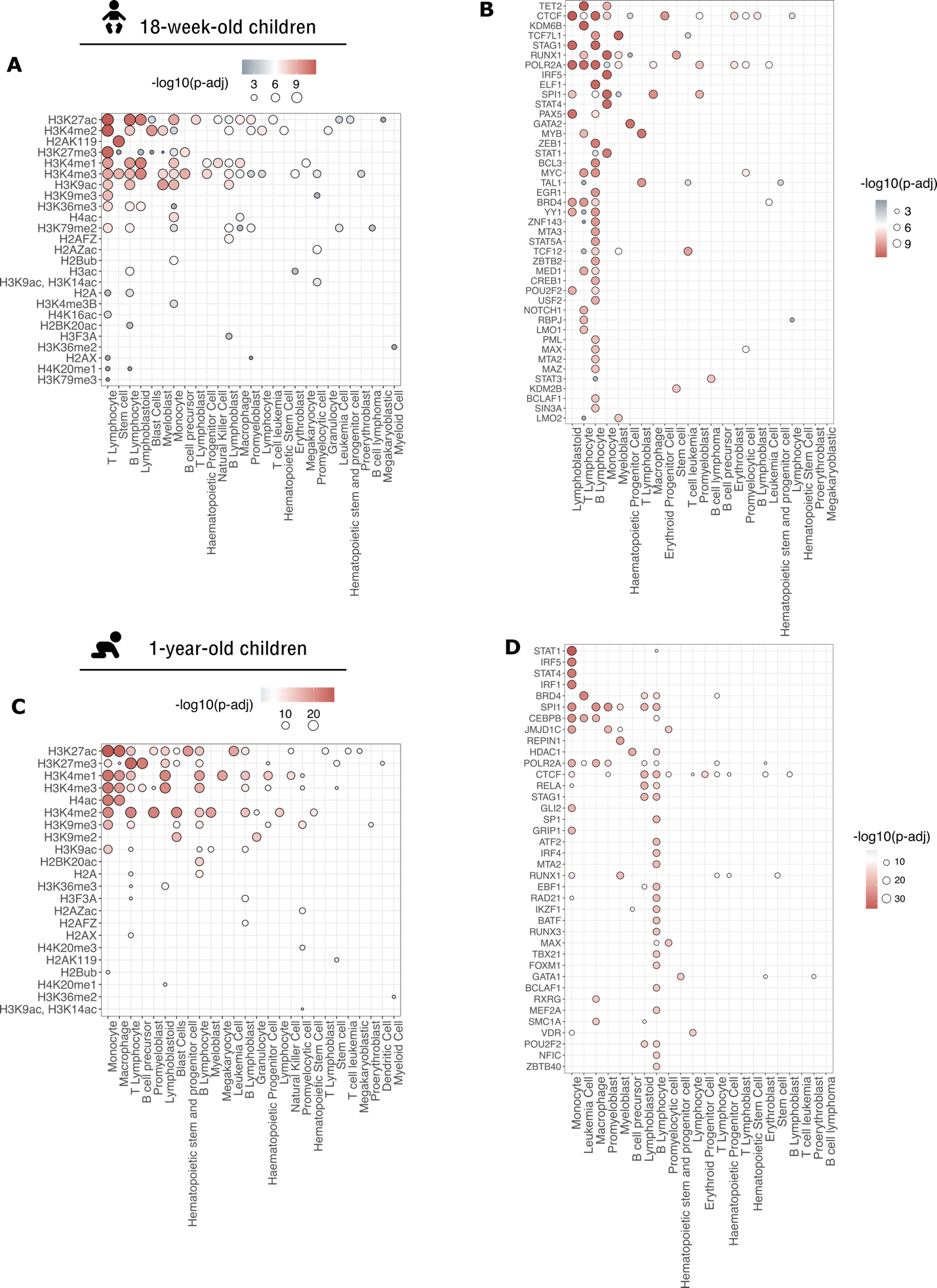

Fig. S7. (A) Histone marks or (B) the top quartile of DNA-binding factors enriched in differential H3K27ac regions of 18-week-old children. Each dot represents the most significant adjusted p-value result for a histone mark (DNA-binding factor) – cell type combination. (C, D) Same as (A,B) but for 1-year old children.

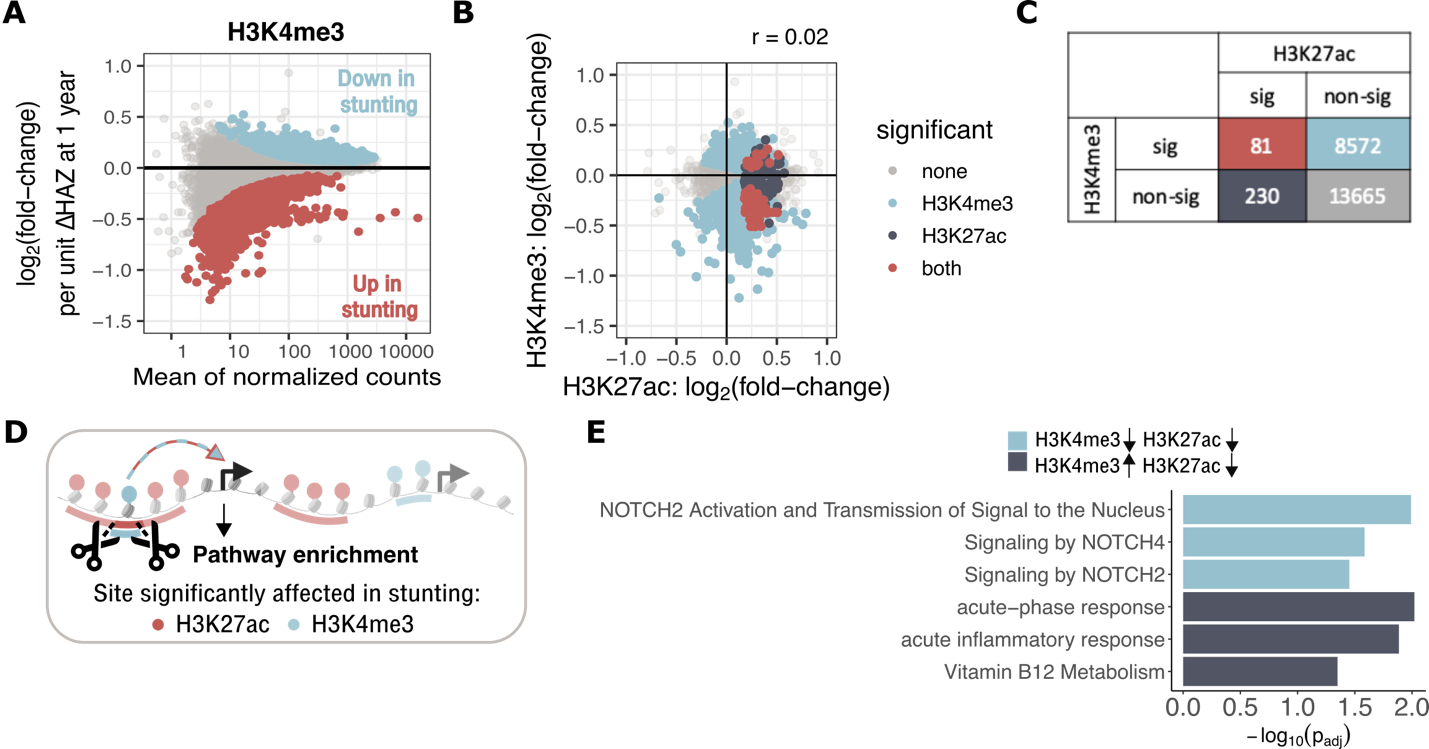

Fig. S8. (A) MA-plot shows H3K4me3 mean coverage of predefined regions and log_2_(fold-change) of coverage with a unit increment of ΔHAZ (1 yr). Colored dots show differential regions with FDR-corrected p-value < 0.05. (B) Scatter plot shows relationship between log_2_(fold-changes) in H3K4me3 vs. H3K27ac with a unit increment of ΔHAZ (1 yr) in 1-year-old children. Each dot represents a region where H3K27ac and H3K4me3 overlap. Grey / light blue / dark blue / red – none / only H3K4me3 / only H3K27ac / both H3K4me3 and H3K27ac were found significantly affected in stunted children. Pearson’s correlation coefficient is mentioned above the plot. (C) Enumeration of regions from (B) with matching color coding. (D) Overview of “high-precision region-centric approach,” where true intersection of H3K27ac and H3K4me3 differential regions are annotated with genes. Compared to Fig. 5f, where the *whole* H3K27ac region is annotated, if it overlaps with H3K4me3. (E) Pathway enrichment of genes associated with intersecting H3K27ac and H3K4me3 regions. Light blue bars show pathways where both H3K2ac and H3K4me3 are downregulated in 1-year old stunted children, dark blue bars pathways in which H3K27ac is downregulated, but H3K4me3 upregulated in stunting.
